# Identifying organ dysfunction trajectory-based subphenotypes in critically ill patients with COVID-19

**DOI:** 10.1101/2020.07.16.20155382

**Authors:** Chang Su, Zhenxing Xu, Katherine Hoffman, Parag Goyal, Monika M Safford, Jerry Lee, Sergio Alvarez-Mulett, Luis Gomez-Escobar, David R Price, John S Harrington, Lisa K Torres, Fernando J Martinez, Thomas R. Campion, Rainu Kaushal, Augustine M.K. Choi, Fei Wang, Edward J. Schenck

**Author notes:** **Author Contributions:** ES and FW for conceptualization, investigation, writing, reviewing and editing of the manuscript. CS for data analysis, drafting, editing and reviewing manuscript. ZX, KH for data analysis. TC for providing data support. PG, MS, SAM, LGE, DP, JS, LT, FM, RK, AC for discussion, commenting and editing the manuscript.

## Abstract

**Rationale:** COVID-19-associated respiratory failure offers the unprecedented opportunity to evaluate the differential host response to a uniform pathogenic insult. Prior studies of Acute Respiratory Distress Syndrome (ARDS) have identified subphenotypes with differential outcomes. Understanding whether there are distinct subphenotypes of severe COVID-19 may offer insight into its pathophysiology.

**Objectives:** To identify and characterize distinct subphenotypes of COVID-19 critical illness defined by the post-intubation trajectory of Sequential Organ Failure Assessment (SOFA) score.

**Methods:** Intubated COVID-19 patients at two hospitals in New York city were leveraged as development and validation cohorts. Patients were grouped into mild, intermediate, and severe strata by their baseline post-intubation SOFA. Hierarchical agglomerative clustering was performed within each stratum to detect subphenotypes based on similarities amongst SOFA score trajectories evaluated by Dynamic Time Warping. Statistical tests defined trajectory subphenotype predictive markers.

**Measurements and Main Results:** Distinct worsening and recovering subphenotypes were identified within each stratum, which had distinct 7-day post-intubation SOFA progression trends. Patients in the worsening suphenotypes had a higher mortality than those in the recovering subphenotypes within each stratum (mild stratum, 29.7% vs. 10.3%, p=0.033; intermediate stratum, 29.3% vs. 8.0%, p=0.002; severe stratum, 53.7% vs. 22.2%, p<0.001). Worsening and recovering subphenotypes were replicated in the validation cohort. Routine laboratory tests, vital signs, and respiratory variables rather than demographics and comorbidities were predictive of the worsening and recovering subphenotypes.

**Conclusions:** There are clear worsening and recovering subphenotypes of COVID-19 respiratory failure after intubation, which are more predictive of outcomes than baseline severity of illness. Organ dysfunction trajectory may be well suited as a surrogate for research in COVID-19 respiratory failure.

**At a Glance Commentary:** *Scientific Knowledge on the Subject:* COVID-19 associated respiratory failure leads to a significant risk of morbidity and mortality. It is clear that there is heterogeneity in the viral-induced host response leading to differential outcomes, even amongst those treated with mechanical ventilation. There are many studies of COVID-19 disease which use intubation status as an outcome or an inclusion criterion. However, there is less understanding of the post intubation course in COVID-19.

*What This Study Adds to the Field:* We have developed and validated a novel subphenotyping model based on post-intubation organ dysfunction trajectory in COVID-19 patients. Specifically, we identified clear worsening and recovering organ dysfunction trajectory subphenotypes, which are more predictive of outcomes than illness severity at baseline. Dynamic inflammatory markers and ventilator variables rather than baseline severity of illness, demographics and comorbidities differentiate the worsening and recovering subphenotypes. Trajectory subphenotypes offer a potential road map for understanding the evolution of critical illness in COVID-19.

## Introduction

The COVID-19 pandemic has created an unprecedented opportunity to explore a large cohort of patients infected with a single pathogen thus providing a window to examine patient variability in response to a uniform insult. Despite this opportunity, distinct subphenotypes of severe-COVID-19 associated respiratory failure remain largely unexplored(1-3). SARS-CoV-2 infection often leads to hypoxemic respiratory failure requiring treatment with mechanical ventilation which meets clinical and pathologic criteria for Acute Respiratory Distress Syndrome (ARDS)(4-6). In COVID-19 respiratory failure, like other forms of ARDS, there is significant risk of morbidity and mortality. However, there is clear heterogeneity in outcomes, even in those treated with mechanical ventilation(4, 5, 7-9). The baseline clinical characteristics and predictors of mortality of those requiring mechanical ventilation have been described(4, 7, 8, 10). These studies offer some insight into a differential host response but are limited to characterizing patients at baseline.

In prior studies of ARDS(11, 12), unique subphenotypes have been described, which identify hyperinflammatory and hypoinflammatory populations with differential demographics, clinical characteristics, inflammatory markers and outcomes. These subphenotypes are primarily characterized by host response inflammatory markers and patterns of organ injury, but are agnostic of the type of insult or infection. In COVID-19, baseline risk stratification may be insufficient to characterize subphenotypes that accurately reflect the complexity of the disease arc(13). Serial, temporally ordered, Sequential Organ Failure Assessment (SOFA)(14-17) and comprehensive Electronic Health Records (EHR) data are well suited to develop data-driven subphenotypes(18), where the goal is to identify coherent patient groups with similar clinical courses. Dynamic time warping (DTW)(19) is a well-established technique for evaluating the similarities among temporal sequences(20, 21). DTW is particularly well suited to evaluate longitudinal changes in organ dysfunction in COVID19. Characterizing a more complete representation of the disease course in COVID19 may offer insight into its pathophysiology.

We used DTW to conduct a two staged post-intubation trajectory analysis of SOFA-based organ dysfunction in patients with COVID19 to identify unique subphenotypes. In order to understand the differential disease course, we then explored clinical and biologic features including demographics, comorbidities, clinical characteristics, inflammatory markers, and treatments predictive of these trajectories.

## Methods

This was a retrospective two staged modeling analysis on two cohorts of intubated COVID-19 patients. The overall workflow of our study is illustrated in Figure 1.

**Figure 1.**
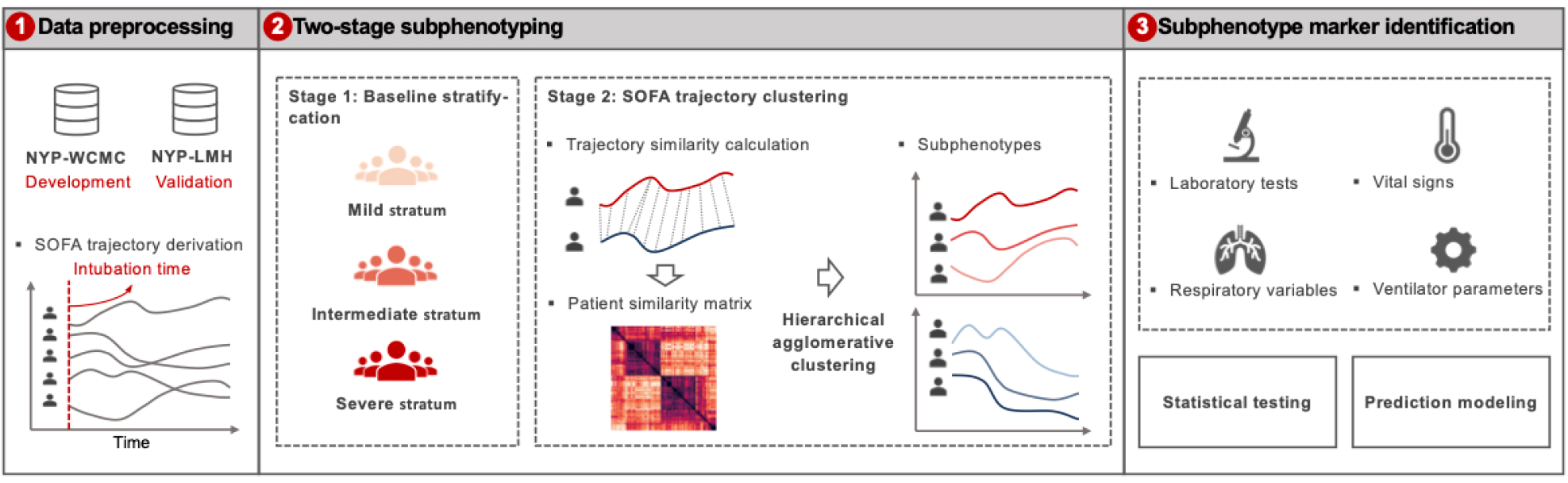
A schematic of the analysis plan. Intubated patients of two cohorts, New NYP-WCMC and NYP-LMH cohorts were analyzed, as development and validation cohorts, respectively. 7-day post-intubation SOFA trajectories were constructed. A two-stage subphenotyping model was then performed on the top of the SOFA trajectories. Statistical testing and prediction modeling were finally performed to identify markers at early-stage after intubation for separating the identified trajectory subphenotypes. Abbreviations: NYP-WCMC=New York Presbyterian Hospital-Weill Cornell Medical Center, NYP-LMH=New York Presbyterian-Lower Manhattan Hospital, SOFA=Sequential Organ Failure Assessment.

### Study design and cohort description

We used individual patient data from two New York Presbyterian (NYP) system hospitals located in New York city: the New York Presbyterian Hospital-Weill Cornell Medical Center (NYP-WCMC), an 862-bed quaternary care hospital, and the New York Presbyterian-Lower Manhattan Hospital (NYP-LMH), a 180-bed non-teaching academic affiliated hospital. Patients were admitted from Mar 3, 2020 to May 12, 2020. SARS-CoV2 diagnosis was made through reverse-transcriptase–PCR assays performed on nasopharyngeal swabs. The critical care response to the pandemic has been previously described(22). The NYP-WCMC cohort was used as the development cohort to derive subphenotypes, and the NYP-LMH cohort was used for validation. The focus of this study was critically ill patients with COVID-19 who were treated with intubation (Supplemental Appendix 1).

### Data collection

We collected all data from either the Weill Cornell-Critical carE Database for Advanced Research (WC-CEDAR), Weill Cornell Medicine COVID Institutional Data Repository (COVID-IDR), or via manual chart abstraction (REDCap). WC-CEDAR aggregates and transforms data from institutional electronic health records for all patients treated in ICUs in NYP-WCMC and NYP-LMH(23). The COVID-IDR contains additional aggregate EHR data on all patients who were tested for SARS-CoV-2 at NYP-WCMC or NYP-LMH. The REDCap database contains high-quality manually abstracted data on all patients who tested positive for COVID-19 at NYP-WCMC or NYP-LMH(24). In our analysis, the patient information incorporated included demographics, laboratory tests, vital signs, and respiratory variables obtained from WC-CEDAR, comorbidity information obtained from the REDCap database, and medication data obtained from the COVID-IDR. Data analyzed were detailed in Supplemental Appendix 2.

### SOFA calculation

The SOFA score is the sum of six organ dysfunction subscores, including cardiovascular, central nervous system (CNS), coagulation, liver, renal, and respiration(14, 17). In this study, the CNS, coagulation, liver, and renal subscores were derived according to the standard SOFA scoring system(14). The respiration subscore was calculated using a combination of the traditional and modified scoring method(25). The cardiovascular SOFA subscore was calculated with additional vasopressors according to a norepinephrine equivalency table, where phenylephrine and vasopressin were converted to a norepinephrine equivalency(26). SOFA scores were derived every 24 hours from the time of intubation, and the worst score within that 24-hour data period was selected for each patient.(14)

### Inclusion exclusion criteria

We included patients with positive results on viral RNA detection by real-time reverse transcriptase polymerase chain reaction (RT-PCR) test from nasopharyngeal swabs specimens and treated with mechanical ventilation at the ICU in NYP-WCM and NYP-LMH. We excluded patients who were less than 18 years old. Since our aim was to identify clinically meaningful organ dysfunction progression patterns of intubated patients, trajectories with low quality (20 (5.7%) patients missing over 50% SOFA records) and outlier trajectories (10 (2.9%) patients with unchanged or heavily fluctuated within the 7-day window after intubation) were excluded from the analysis (Supplemental Appendix 3 and Figure E-1).

### Subphenotyping model description

SOFA scores were derived every 24 hours and post intubation 7 day SOFA trajectories were constructed for analysis. Missing values within a trajectory were imputed based on the last observation carried forward (LOCF) strategy.

A two-staged subphenotyping method was performed to derive SOFA trajectory subphenotypes (Figure 1). In the first stage, we used baseline SOFA to group patients with a similar upfront risk of death(17), as additive organ dysfunction has previously been identified to be associated with poor outcomes in COVID19(8). We partitioned the patients into three baseline severity strata (mild, intermediate, and severe) according to their SOFA scores within the first 24 hours after intubation. The SOFA score cut-offs were set to 0-10, 11-12, and 13-24 in order to obtain a balanced distribution of patients across the three strata. In the second stage, we identified the subphenotypes with similar 7-day SOFA progression patterns. Dynamic Time Warping (DTW)(19) was adopted to evaluate the similarities between pairwise patient SOFA trajectories within each baseline stratum and then hierarchical agglomerative clustering (HAC)(27) was performed on these similarities to derive the similar patient clusters as trajectory subphenotypes. DTW can account for the differences among the evolution heterogeneity among the temporal curves and is thus able to evaluate their similarity more robustly.(19) The optimal numbers of subphenotypes were determined by clear separation illustrated by clustergram according to the McClain index(28). DTW was implemented with Python 3.7 based on tslearn package 0.3.1 and HAC was performed with Python 3.7 based on scikit-learn package 0.22.2.

To validate these findings, we replicated these subphenotypes from the NYP-LMH cohort.

### Clinical outcomes

We analyzed 30-day all-cause mortality as the primary outcome for patients within each phenotype. Successful extubation or need for tracheostomy within 30 days after intubation were secondary outcomes.

### Statistical analysis

We examined the associations between patient characteristics and clinical variables and the identified trajectory subphenotypes, to see if there are early markers that can discriminate between them. Patient characteristics we investigated included demographics, comorbidities, medications prescribed within the window from 3-day before to 5-day after intubation, and blood type(29). Laboratory test results included: complete blood count, basic metabolic panel, liver function tests, coagulation profile and inflammatory markers including d-dimer, fibrinogen, ferritin, erythrocyte sedimentation rate, lactic acid, troponin, lactate dehydrogenase, creatine kinase, procalcitonin and C-reactive protein. Vital signs included: GCS, mean arterial pressure and temperature, urine output. Respiratory variables included: P/F ratio, FiO2, Pao2, PaCO2, PH, PEEP, peak inspiratory pressure, plateau pressure, driving pressure, static compliance, minute ventilation, ventilatory ratio, and tidal volume indexed to ideal body weight at day 1 and day 3 post-intubation.

Univariate statistical tests were performed in those association analyses. Specifically, one-way analysis of variance (ANOVA, with Tukey HSD post hoc test), Kruskal–Wallis test (with Dunn post hoc test), student’s t-test, Mann-Whitney test, Chi-square test, and Fisher’s exact test have been used whenever appropriate. The p-values were then corrected for multiple testing using false discovery rate (FDR) estimation. Analysis of covariance (ANCOVA) for the between-strata/subphenotypes comparisons was also applied based on the generalized linear model (GLM) with adjustment on age at baseline. All statistical tests were performed with Python 3.7 based on statsmodels package 0.11.1.

### Subphenotype prediction modeling

We trained a random forest model with the trajectory subphenotypes as targets and the patient clinical characteristics at specific time points after intubation as input predictors to define if these trajectory subphenotypes can be predicted early. Our implementation was with Python 3.7 based on scikit-learn package 0.22.2. Candidate predictors included demographics, comorbidities, medications prescribed around the intubation event, SOFA subscores, laboratory tests, vital signs, and respiratory variables as described above. Prediction performances were measured by area under the receiver operating characteristics (AUC-ROC). The importance of predictors was visualized as a heatmap to demonstrate their contributions on subphenotype prediction.

### IRB approval

The study is approved by the IRB of Weill Cornell Medicine with protocol number 20-04021909.

## Results

### Patients and baseline severity strata

A total of 318 mechanically ventilated COVID-19 patients from the NYP-WCMC cohort were included for analysis, consisting of 100 females (31.45%) and an average age of 62.78 ± 14.34. One day post-intubation the mean SOFA score for this cohort is 11.89 ± 2.56. A total of 84 mechanically ventilated COVID-19 patients from the NYP-LMH were included as a validation cohort, consisting of 33 (39.29%) females and an average age of 66.06 ± 13.06. One day post-intubation the mean SOFA score is 12.51 ± 2.25. The clinical characteristics of both cohorts are summarized in Table 1.

**Table 1.** Clinical characteristics of the studied cohorts.

| Variable | NYP-WCMC cohort |  |  |  | NYP-LMH validation cohort |  |  |  |
| --- | --- | --- | --- | --- | --- | --- | --- | --- |
|  | All | Mild stratum | Intermediate stratum | Severe stratum | All | Mild stratum | Intermediate stratum | Severe stratum |
| # of patients (%) | 318 | 76 (23.90) | 116 (36.48) | 126 (39.62) | 84 | 10 (11.90) | 35 (41.67) | 39 (46.43) |
| **Demographics** |  |  |  |  |  |  |  |  |
| Age, Mean (SD) | 62.78 (14.34) | 61.47 (16.51) | 60.53 (14.14) | 65.64 (12.52) | 66.06 (13.06) | 61.00 (17.10) | 61.63 (11.46) | 71.33 (11.07) |
| Sex female, n (%) | 100 (31.45%) | 23 (30.26%) | 38 (32.76%) | 39 (30.95%) | 33 (39.29%) | 4 (40.00%) | 19 (54.29%) | 10 (25.64%) |
| CAUCASIAN, n (%) | 91 (28.62%) | 20 (26.32%) | 39 (33.62%) | 32 (25.40%) | 7 (8.33%) | 0 (0.00%) | 4 (11.43%) | 3 (7.69%) |
| AFRICAN AMERICAN, n (%) | 27 (8.49%) | 3 (3.95%) | 5 (4.31%) | 19 (15.08%) | 7 (8.33%) | 0 (0.00%) | 0 (0.00%) | 7 (17.95%) |
| ASIAN/PACIFIC ISLANDER, n (%) | 33 (10.38%) | 11 (14.47%) | 9 (7.76%) | 13 (10.32%) | 32 (38.10%) | 5 (50.00%) | 12 (34.29%) | 15 (38.46%) |
| MULTI-RACIAL, n (%) | 86 (27.04%) | 21 (27.63%) | 34 (29.31%) | 31 (24.60%) | 10 (11.90%) | 2 (20.00%) | 5 (14.29%) | 3 (7.69%) |
| BMI, Mean (SD) | 29.53 (8.40) | 29.23 (9.06) | 30.75 (9.17) | 28.59 (7.01) | 28.70 (7.70) | 26.67 (3.94) | 30.03 (9.94) | 28.03 (5.61) |
| **Comorbidities** |  |  |  |  |  |  |  |  |
| Coronary Artery Disease, n (%) | 49 (15.41%) | 7 (9.21%) | 17 (14.66%) | 25 (19.84%) | 11 (13.10%) | 1 (10.00%) | 1 (2.86%) | 9 (23.08%) |
| Cerebrovascular accident (Stroke), n (%) | 20 (6.29%) | 3 (3.95%) | 7 (6.03%) | 10 (7.94%) | 4 (4.76%) | 0 (0.00%) | 0 (0.00%) | 4 (10.26%) |
| Heart Failure, n (%) | 21 (6.60%) | 3 (3.95%) | 9 (7.76%) | 9 (7.14%) | 3 (3.57%) | 0 (0.00%) | 1 (2.86%) | 2 (5.13%) |
| Hypertension, n (%) | 167 (52.52%) | 35 (46.05%) | 57 (49.14%) | 75 (59.52%) | 50 (59.52%) | 5 (50.00%) | 17 (48.57%) | 28 (71.79%) |
| Diabetes Mellitus, n (%) | 94 (29.56%) | 17 (22.37%) | 30 (25.86%) | 47 (37.30%) | 35 (41.67%) | 4 (40.00%) | 12 (34.29%) | 19 (48.72%) |
| Pulmonary Disease, n (%) | 63 (19.81%) | 15 (19.74%) | 22 (18.97%) | 26 (20.63%) | 15 (17.86%) | 2 (20.00%) | 4 (11.43%) | 9 (23.08%) |
| Renal Disease, n (%) | 26 (8.18%) | 5 (6.58%) | 5 (4.31%) | 16 (12.70%) | 7 (8.33%) | 0 (0.00%) | 2 (5.71%) | 5 (12.82%) |
| Cirrhosis, n (%) | 5 (1.57%) | 3 (3.95%) | 0 (0.00%) | 2 (1.59%) | 1 (1.19%) | 0 (0.00%) | 0 (0.00%) | 1 (2.56%) |
| Hepatitis, n (%) | 4 (1.26%) | 1 (1.32%) | 0 (0.00%) | 3 (2.38%) | 2 (2.38%) | 0 (0.00%) | 1 (2.86%) | 1 (2.56%) |
| HIV, n (%) | 4 (1.26%) | 1 (1.32%) | 2 (1.72%) | 1 (0.79%) | 1 (1.19%) | 0 (0.00%) | 1 (2.86%) | 0 (0.00%) |
| Active Cancer, n (%) | 21 (6.60%) | 3 (3.95%) | 2 (1.72%) | 16 (12.70%) | 2 (2.38%) | 0 (0.00%) | 0 (0.00%) | 2 (5.13%) |
| Transplant, n (%) | 14 (4.40%) | 5 (6.58%) | 3 (2.59%) | 6 (4.76%) | 1 (1.19%) | 0 (0.00%) | 0 (0.00%) | 1 (2.56%) |
| Inflammatory Bowel Disease, n (%) | 7 (2.20%) | 2 (2.63%) | 2 (1.72%) | 3 (2.38%) | 0 (0.00%) | 0 (0.00%) | 0 (0.00%) | 0 (0.00%) |
| Rheumatologic Disease, n (%) | 15 (4.72%) | 4 (5.26%) | 3 (2.59%) | 8 (6.35%) | 3 (3.57%) | 0 (0.00%) | 2 (5.71%) | 1 (2.56%) |
| Other Immunosuppressed State, n (%) | 12 (3.77%) | 4 (5.26%) | 1 (0.86%) | 7 (5.56%) | 0 (0.00%) | 0 (0.00%) | 0 (0.00%) | 0 (0.00%) |
| **Baseline SOFA scores** |  |  |  |  |  |  |  |  |
| Cardiovascular, Mean (SD) | 3.02 (1.35) | 1.32 (1.34) | 3.41 (0.88) | 3.69 (0.70) | 3.45 (1.03) | 1.40 (1.02) | 3.57 (0.80) | 3.87 (0.40) |
| Central nervous system, Mean (SD) | 3.72 (0.68) | 3.34 (1.13) | 3.72 (0.47) | 3.94 (0.24) | 3.39 (0.74) | 2.60 (1.36) | 3.37 (0.48) | 3.62 (0.54) |
| Coagulation, Mean (SD) | 0.15 (0.47) | 0.12 (0.40) | 0.04 (0.20) | 0.28 (0.64) | 0.13 (0.40) | 0.00 (0.00) | 0.11 (0.40) | 0.18 (0.45) |
| Liver, Mean (SD) | 0.24 (0.56) | 0.20 (0.46) | 0.14 (0.43) | 0.37 (0.67) | 0.20 (0.48) | 0.10 (0.30) | 0.14 (0.42) | 0.28 (0.55) |
| Renal, Mean (SD) | 0.94 (1.32) | 0.16 (0.54) | 0.35 (0.67) | 1.96 (1.44) | 1.36 (1.35) | 0.50 (0.67) | 0.37 (0.64) | 2.46 (1.08) |
| Respiration, Mean (SD) | 3.81 (0.58) | 3.45 (0.89) | 3.89 (0.45) | 3.97 (0.25) | 3.98 (0.22) | 4.00 (0.00) | 3.94 (0.33) | 4.00 (0.00) |
| SOFA score, Mean (SD) | 11.89 (2.56) | 8.58 (1.84) | 11.55 (0.58) | 14.20 (1.46) | 12.51 (2.25) | 8.60 (2.11) | 11.51 (0.50) | 14.41 (1.08) |
| **30-day Clinical Outcomes** |  |  |  |  |  |  |  |  |
| Extubation, n (%) | 138 (43.40%) | 40 (52.63%) | 54 (46.55%) | 44 (34.92%) | 31 (36.90%) | 2 (20.00%) | 15 (42.86%) | 14 (35.90%) |
| Mortality, n (%) | 77 (24.21%) | 14 (18.42%) | 18 (15.52%) | 45 (35.71%) | 26 (30.95%) | 4 (40.00%) | 8 (22.86%) | 14 (35.90%) |
| Tracheostomy, n (%) | 41 (12.89%) | 10 (13.16%) | 18 (15.52%) | 13 (10.32%) | 6 (7.14%) | 0 (0.00%) | 3 (8.57%) | 3 (7.69%) |

Abbreviation: BMI=body mass index, HIV=Human Immunodeficiency Virus, NYP-WCMC=New York Presbyterian Hospital-Weill Cornell Medical Center, NYP-LMH=New York Presbyterian-Lower Manhattan Hospital, SD=standard deviation, SOFA=Sequential Organ Failure Assessment

For the NYP-WCMC cohort, patients were first partitioned into mild, intermediate, and severe strata based on the SOFA scores within one day after intubation, consisting of 76 (23.29%), 116 (36.48%), and 126 (39.62%) patients, respectively; while for the NYP-LMH validation cohort, the three strata consist of 10 (11.90%), 35 (41.67%), and 39 (46.43%) patients, respectively. As shown in Table 1, the patients in both NYP-WCMC and NYP-LMH cohorts exhibit additive patterns of post intubation baseline organ dysfunction according to the SOFA subscores. Specifically, CNS and respiration dysfunction were present in the mild stratum; the intermediate stratum had additional cardiovascular dysfunction on top of CNS and respiratory dysfunction compared to the mild stratum; and the severe stratum had renal dysfunction in addition to all other organ failure. Liver and coagulation dysfunction were rare in all strata. Patients in the severe stratum were generally older and were more likely to suffer from chronic comorbidities at baseline.

### SOFA trajectory subphenotypes

The clustergrams built upon the pairwise SOFA trajectory distance matrix derived by DTW are shown in Supplemental Figure E-2. The optimal number of subphenotypes within each stratum as determined by the McClain Index(28) are shown in Supplemental Table E-1, suggesting two being the best choice across all strata in both cohorts. Figure 2 demonstrates the individual averaged SOFA curves for patients in the two subphenotypes across all strata: a worsening subphenotype of which SOFA score increased within the 7-day observation window, and a recovering subphenotype of which SOFA score improved. The clinical characteristics of these subphenotypes were summarized in Table 2. Overall, there was no marked difference in terms of demographics, comorbidity burden, and pattern of organ dysfunction (distribution of SOFA subscores and total score) between the worsening and recovering subphenotypes within each baseline severity stratum at baseline. This suggests that, though the subphenotypes varied in 7-day organ dysfunction progression patterns, they have similar clinical status immediately after intubation. We further investigated medications prescribed within each subphenotype and didn’t find significant signal as well (Supplemental Table E-3). In addition, clinical characteristics and medications of the subphenotypes re-derived in the NYP-LMH validation cohort were summarized in Supplemental Tables E-2 and 4.

**Table 2.** Clinical characteristics of the trajectory subphenotypes in NYP-WCMC cohort.

| Variable | Mild stratum<br>(SOFA 0-10, n=76) |  |  | Intermediate stratum<br>(SOFA 11-12, n=116) |  |  | Severe stratum<br>(SOFA 13-24, n=126) |  |  |
| --- | --- | --- | --- | --- | --- | --- | --- | --- | --- |
|  | Worsening | Recovering | p-value<sup>†</sup> | Worsening | Recovering | p-value<sup>†</sup> | Worsening | Recovering | p-value<sup>†</sup> |
| **Total #** | **37** | **39** | **-** | **41** | **75** | **-** | **54** | **72** | **-** |
| **Demographics** |  |  |  |  |  |  |  |  |  |
| Age, Mean (SD) | 61.08 (14.95) | 61.85 (17.86) | 0.240 | 63.80 (13.90) | 58.73 (13.95) | 0.059 | 65.72 (11.05) | 65.58 (13.52) | 0.951 |
| Sex female, n (%) | 9 (24.32%) | 14 (35.90%) | 0.323 | 13 (31.71%) | 25 (33.33%) | 1.000 | 17 (31.48%) | 22 (30.56%) | 1.000 |
| CAUCASIAN, n (%) | 9 (24.32%) | 11 (28.21%) |  | 14 (34.15%) | 25 (33.33%) |  | 16 (29.63%) | 16 (22.22%) |  |
| AFRICAN AMERICAN, n (%) | 1 (2.70%) | 2 (5.13%) | 0.927 | 2 (4.88%) | 3 (4.00%) | 0.883 | 8 (14.81%) | 11 (15.28%) | 0.846 |
| ASIAN/PACIFIC ISLANDER, n (%) | 5 (13.51%) | 6 (15.38%) |  | 3 (7.32%) | 6 (8.00%) |  | 4 (7.41%) | 9 (12.50%) |  |
| MULTI-RACIAL, n (%) | 12 (32.43%) | 9 (23.08%) |  | 10 (24.39%) | 24 (32.00%) |  | 13 (24.07%) | 18 (25.00%) |  |
| BMI, Mean (SD) | 29.42 (10.01) | 29.07 (8.21) | 0.435 | 29.99 (7.25) | 31.18 (10.09) | 0.416 | 29.79 (7.01) | 27.71 (6.89) | 0.018 |
| **Comorbidities** |  |  |  |  |  |  |  |  |  |
| Coronary Artery Disease, n (%) | 5 (13.51%) | 2 (5.13%) | 0.248 | 5 (13.51%) | 2 (5.13%) | 0.248 | 11 (20.37%) | 14 (19.44%) | 0.824 |
| Cerebrovascular accident (Stroke), n (%) | 0 (0.00%) | 3 (7.69%) | 0.241 | 0 (0.00%) | 3 (7.69%) | 0.241 | 2 (3.70%) | 8 (11.11%) | 0.189 |
| Heart Failure, n (%) | 2 (5.41%) | 1 (2.56%) | 0.604 | 2 (5.41%) | 1 (2.56%) | 0.604 | 4 (7.41%) | 5 (6.94%) | 1.000 |
| Hypertension, n (%) | 15 (40.54%) | 20 (51.28%) | 0.479 | 15 (40.54%) | 20 (51.28%) | 0.479 | 35 (64.81%) | 40 (55.56%) | 0.248 |
| Diabetes Mellitus, n (%) | 6 (16.22%) | 11 (28.21%) | 0.275 | 6 (16.22%) | 11 (28.21%) | 0.275 | 24 (44.44%) | 23 (31.94%) | 0.130 |
| Pulmonary Disease, n (%) | 7 (18.92%) | 8 (20.51%) | 1.000 | 7 (18.92%) | 8 (20.51%) | 1.000 | 14 (25.93%) | 12 (16.67%) | 0.184 |
| Renal Disease, n (%) | 0 (0.00%) | 5 (12.82%) | 0.055 | 0 (0.00%) | 5 (12.82%) | 0.055 | 8 (14.81%) | 8 (11.11%) | 0.589 |
| Cirrhosis, n (%) | 1 (2.70%) | 2 (5.13%) | 1.000 | 1 (2.70%) | 2 (5.13%) | 1.000 | 1 (1.85%) | 1 (1.39%) | 1.000 |
| Hepatitis, n (%) | 0 (0.00%) | 1 (2.56%) | 1.000 | 0 (0.00%) | 1 (2.56%) | 1.000 | 2 (3.70%) | 1 (1.39%) | 0.572 |
| HIV, n (%) | 0 (0.00%) | 1 (2.56%) | 1.000 | 0 (0.00%) | 1 (2.56%) | 1.000 | 0 (0.00%) | 1 (1.39%) | 1.000 |
| Active Cancer, n (%) | 2 (5.41%) | 1 (2.56%) | 0.604 | 2 (5.41%) | 1 (2.56%) | 0.604 | 10 (18.52%) | 6 (8.33%) | 0.102 |
| Transplant, n (%) | 1 (2.70%) | 4 (10.26%) | 0.359 | 1 (2.70%) | 4 (10.26%) | 0.359 | 5 (9.26%) | 1 (1.39%) | 0.081 |
| Inflammatory Bowel Disease, n (%) | 0 (0.00%) | 2 (5.13%) | 0.494 | 0 (0.00%) | 2 (5.13%) | 0.494 | 1 (1.85%) | 2 (2.78%) | 1.000 |
| Rheumatologic Disease, n (%) | 0 (0.00%) | 4 (10.26%) | 0.116 | 0 (0.00%) | 4 (10.26%) | 0.116 | 3 (5.56%) | 5 (6.94%) | 1.000 |
| Other Immunosuppressed State, n (%) | 2 (5.41%) | 2 (5.13%) | 1.000 | 2 (5.41%) | 2 (5.13%) | 1.000 | 5 (9.26%) | 2 (2.78%) | 0.129 |
| **Baseline SOFA scores** |  |  |  |  |  |  |  |  |  |
| Cardiovascular, Mean (SD) | 1.41 (1.26) | 1.23 (1.40) | 0.220 | 3.27 (0.86) | 3.48 (0.88) | 0.061 | 3.65 (0.72) | 3.72 (0.67) | 0.286 |
| Central nervous system, Mean (SD) | 3.41 (1.03) | 3.28 (1.22) | 0.358 | 3.71 (0.45) | 3.73 (0.47) | 0.342 | 3.94 (0.23) | 3.93 (0.25) | 0.379 |
| Coagulation, Mean (SD) | 0.05 (0.32) | 0.18 (0.45) | 0.033 | 0.05 (0.22) | 0.04 (0.20) | 0.415 | 0.31 (0.74) | 0.25 (0.55) | 0.499 |
| Liver, Mean (SD) | 0.27 (0.50) | 0.13 (0.40) | 0.059 | 0.17 (0.44) | 0.12 (0.43) | 0.148 | 0.37 (0.75) | 0.36 (0.61) | 0.369 |
| Renal, Mean (SD) | 0.24 (0.67) | 0.08 (0.35) | 0.103 | 0.46 (0.63) | 0.29 (0.69) | 0.023 | 1.94 (1.45) | 1.97 (1.44) | 0.466 |
| Respiration, Mean (SD) | 3.68 (0.70) | 3.23 (1.00) | 0.021 | 3.85 (0.52) | 3.91 (0.41) | 0.330 | 3.93 (0.38) | 4.00 (0.00) | 0.052 |
| SOFA score, Mean (SD) | 9.05 (1.45) | 8.13 (2.04) | 0.009 | 11.51 (0.50) | 11.57 (0.61) | 0.164 | 14.15 (1.57) | 14.24 (1.37) | 0.253 |

<sup>†</sup> p-value calculated by Chi-square test/Fisher's exact test, or student's t-test/Mann-Whitney test where appropriate.
\*\* False discovery rate corrected p-value < 0.05
Abbreviation: BMI=body mass index, HIV=Human Immunodeficiency Virus, NYP-WCMC=New York Presbyterian Hospital-Weill Cornell Medical Center, SD=standard deviation, SOFA=Sequential Organ Failure Assessment

**Figure 2.**
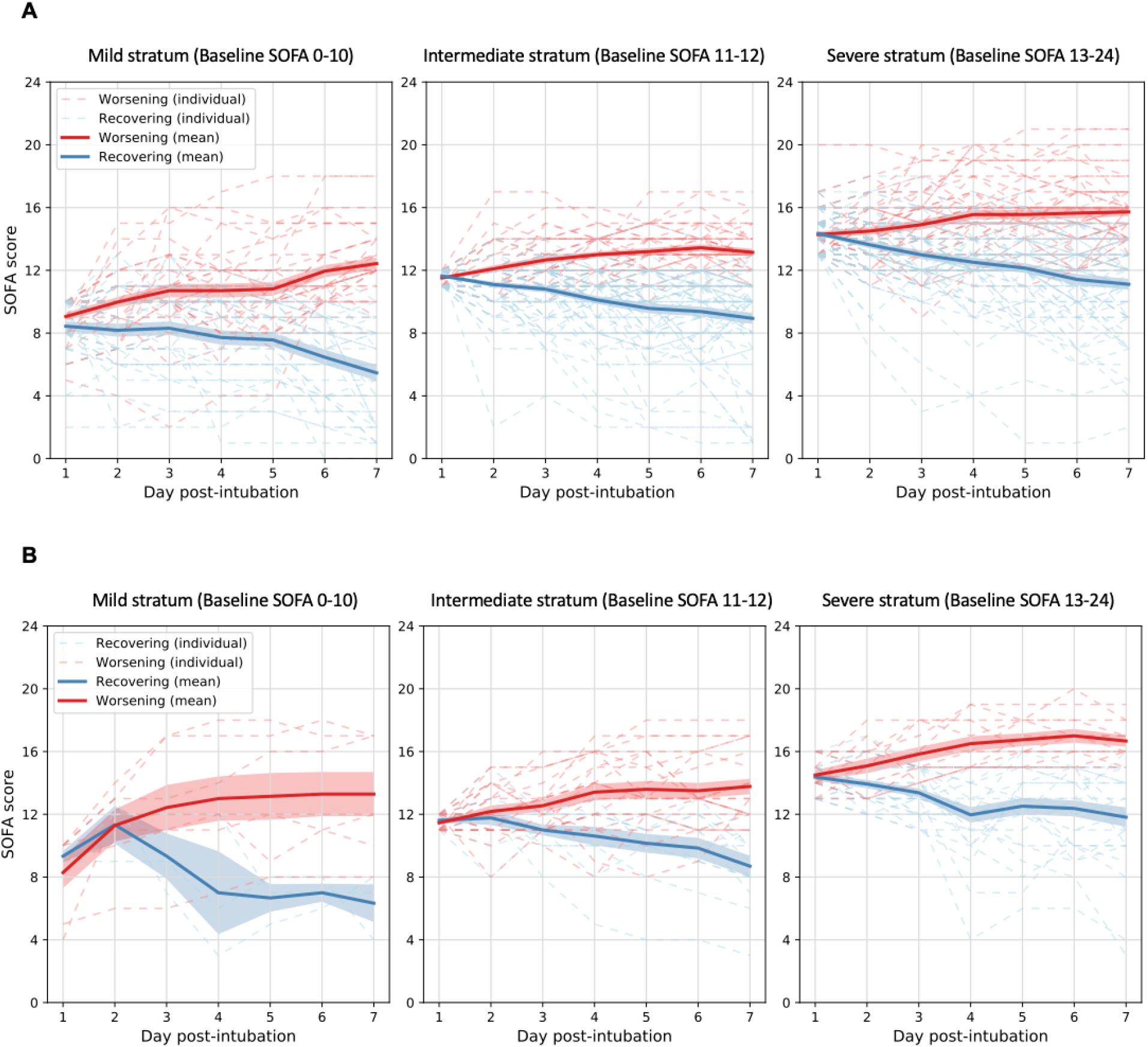
Averaged Sequential Organ Failure Assessment (SOFA) trajectories of the identified subphenotypes. Solid curves are mean SOFA trajectories of the subphenotypes, while dashed curves are individual SOFA trajectories of the patients. *(A)* SOFA trajectories of subphenotypes derived in NYP-WCMC cohort. *(B)* SOFA trajectories of subphenotypes derived in NYP-LMH validation cohort.

### 30-days clinical outcomes

Statistics of 30-day post-intubation clinical primary and secondary outcomes (mortality, extubation, and tracheostomy) of subphenotypes were illustrated in Figure 2A and Supplemental Figure E-3A. The worsening subphenotypes, across baseline strata, suffered from a significantly higher risk of mortality within the 30-day window after intubation (worsening vs recovering, mortality proportion: mild stratum, 29.7% vs. 10.3%, p=0.033; intermediate stratum, 29.3% vs. 8.0%, p=0.002; severe stratum, 53.7% vs. 22.2%, p<0.001). The recovering subphenotypes, across all baseline strata, showed significantly higher extubation proportions within the 30-day window compared to the worsening subphenotypes (recovering vs. worsening, extubation proportion: mild stratum, 76.9% vs. 27.0%, p<0.001; intermediate stratum, 54.7% vs. 31.7%, p=0.018; severe stratum 50.0% vs. 14.8%, p<0.001). There was no significant difference of 30-day tracheostomy detected between the subphenotypes. Importantly, the recovering subphenotype within the severe baseline stratum had a lower mortality risk compared to the worsening subphenotypes at mild and intermediate baseline strata.

The trajectory subphenotypes derived in the NYP-LMH validation cohort had similar trends in all three clinical outcomes within the 30-day window after intubation (see Figure 2A and Supplemental Figure E-3B). Across all baseline strata, the worsening subphenotypes accounted for higher risk of mortality (worsening vs recovering, mortality proportion: mild stratum, 57.1% vs. 0.0%, p=0.200; intermediate stratum, 31.8% vs. 7.7%, p=0.211; severe stratum, 83.3% vs. 17.4%, p<0.001), while the recovering subphenotypes showed higher extubation proportion within 30-days after intubation (recovering vs. worsening, extubation proportion: mild stratum, 33.3% vs. 14.3%, p=0.490; intermediate stratum, 69.2% vs. 27.3%, p=0.015; severe stratum, 48.1% vs. 9.1%, p=0.017).

### Correlation of subphenotypes with early-stage markers

Vital signs, laboratory variables, and respiratory variables were evaluated to identify early-stage markers predictive of the two-stage classification. First of all, the three baseline strata of the NYP-WCMC cohort were observed to be well separated by a series of clinical variables in addition to the differential organ dysfunction pattern noted above (Supplemental Table E-5). For instance, ANOVA(or Kruskal–Wallis test) results showed significantly increased laboratory values like procalcitonin, ferritin, lactate dehydrogenase (LDH), and creatinine, and decreased carbon dioxide within the severe stratum, at day 1 post-intubation; Vital sign such as the Glasgow Coma Scale (GCS) and urine output volume and respiratory variables such as peak inspiratory pressure (PIP) were also observed to separate the strata; the mild stratum was associated with significantly decreased levels of troponin, creatinine, and glucose. Detailed statistical analyses are described in Supplemental Table E-5. Statistics of these clinical variables across baseline strata within the NYP-LMH validation cohort showed similar signals and were detailed in Supplemental Table E-6.

We further evaluated the clinical variable values at day 1 and day 3 post-intubation to compare the worsening and recovering subphenotypes within each baseline stratum. Markers, which significantly separated the worsening and recovering subphenotypes (p-value<0.05), varied across different baseline strata (Supplemental Table E-7). Specifically, on day 1, laboratory tests including white blood cell (WBC) count, procalcitonin, CRP, creatinine, neutrophil count, and globulin, and respiratory variables such as minute ventilation and ratio of tidal volume to predicted body weight (PBW) were found to significantly differentiate the trajectory subphenotypes within the mild stratum. Markers that significantly separated the trajectory subphenotypes within the intermediate stratum on day 1 included AST, creatinine, LDH and FiO2. The severe stratum had more markers separating the worsening and recovering subphenotypes on day 1, including laboratory tests like lymphocyte count, platelet count, triglycerides, procalcitonin level, ferritin, troponin, LDH, creatinine, and AST, respiratory variables such as PIP, PaO2, PaO2/FiO2 ratio (P/F ratio), and SpO2. In addition, more markers separating subphenotypes emerged within each stratum on day 3 after intubation. For instance, additional day 3 markers including GCS, platelet, sodium, and glucose were found within the mild stratum. For the intermediate stratum, GCS, albumin, WBC count, lymphocyte percentage, CRP, sodium, neutrophil percentage and count, carbon dioxide level, urine output volume, and P/F ratio were found to be additional markers on day 3. The additional markers found within the severe stratum on day 3 included vital signs GCS and temperature, lymphocyte percentage, hemoglobin level, albumin level, potassium level, ESR, CK, carbon dioxide level, and lactic acid, positive end-expiratory pressure (PEEP), plateau pressure, minute ventilation, static compliance, driving pressure, FiO2, and ventilator ratio.

As shown in Supplemental Table E-8, most markers identified within the NYP-WCMC cohort showed consistent signals within the NYP-LMH subphenotypes, even though some significance vanishes, as the confidence intervals were wide.

### Subphenotype prediction models

We trained random forest models for predicting the worsening and recovering trajectory subphenotypes within each baseline stratum according to the early stage marker values. Overall, as shown in Supplemental Figure E-4, within the mild, intermediate, and severe strata, the prediction models achieved the AUC-ROCs of 0.70 (95% confidence interval [CI], [0.59, 0.80]), 0.67 (95% CI, [0.62, 0.72]), and 0.73 (95% CI, [0.67, 0.79]) respectively, with the predictor values evaluated at day 1 post-intubation. AUC-ROCs of the models increased to 0.77 (95% CI, [0.66, 0.89]), 0.77 (95% CI, [0.74, 0.81]), and 0.80 (95% CI, [0.75, 0.85]), with the predictor values evaluated at day 3 post-intubation; and to 0.83 (95% CI, [0.72, 0.94]), 0.91 (95% CI, [0.88, 0.94]), and 0.88 (95% CI, [0.80, 0.95]), with the predictor values evaluated at day 5 post-intubation.

Importance of the predictors were illustrated as heatmaps, where color intensity represents the normalized importance of specific predictors (Supplemental Figure E-5). Generally, predictor importance varied as the progress of time. Models trained on day 1-3 after intubation were observed to involve more contributions from the laboratory tests, vital signs, respiratory variables than other predictors; SOFA subscores, especially cardiovascular, CNS, and renal subscores showed relatively higher importance over models trained on day 4 or 5 data within the intermediate and severe strata. Age contributed to day 1-3 prediction to some extent, while other demographics, medications and comorbidities showed weak importance in prediction.

### Correlation to blood type

We finally assessed ABO/RH blood type distribution across the subphenotypes (Supplemental Tables E-9 and 10). Overall, there is no significant signal detected.

## Discussion

In this study, we identified novel trajectory subphenotypes of COVID-19 patients with an objective machine learning approach. The subphenotypes we identified are based on organ dysfunction trajectory over 7-days following intubation, which is different from existing data-driven subphenotyping methods that focus on patient data at a specific timestamp (12, 30, 31). The use of novel methodology, in addition to the robust size of our cohort, ensure that the identified trajectory based subphenotypes are less likely to suffer from cognitive bias(13) and are likely to be temporally stable(32). More concretely, we adopted a divide and conquer approach to identify the subphenotypes. Prior research has identified that additive organ dysfunction is predictive of increased mortality in COVID-19 associated ARDS(8). Therefore, we divided the patients into three different baseline strata (mild, intermediate and severe) according to additive SOFA based organ dysfunction. We identified two salient trajectory subphenotypes within each stratum.

Importantly, the baseline demographics, comorbidities and pattern of organ dysfunction did not differ between the worsening and recovering subphenotypes at each stratum. This suggests the existence of differential progression pathways that are irrespective of baseline risk factors for severe disease. This finding is unique compared to other subphenotyping projects as we are including a more complete picture of the disease course(12, 30, 31). It also highlights the temporal heterogeneity of COVID-19 and the importance of avoiding prognostication based on early post intubation clinical characteristics. We found that the worsening subphenotypes in the baseline mild and intermediate strata showed an even higher risk of death compared to the recovering subphenotype within the baseline severe stratum (Figure 3). Indeed, there is an urgent need to understand the pathophysiology of progressive non-pulmonary organ dysfunction in this disease.

**Figure 3.**
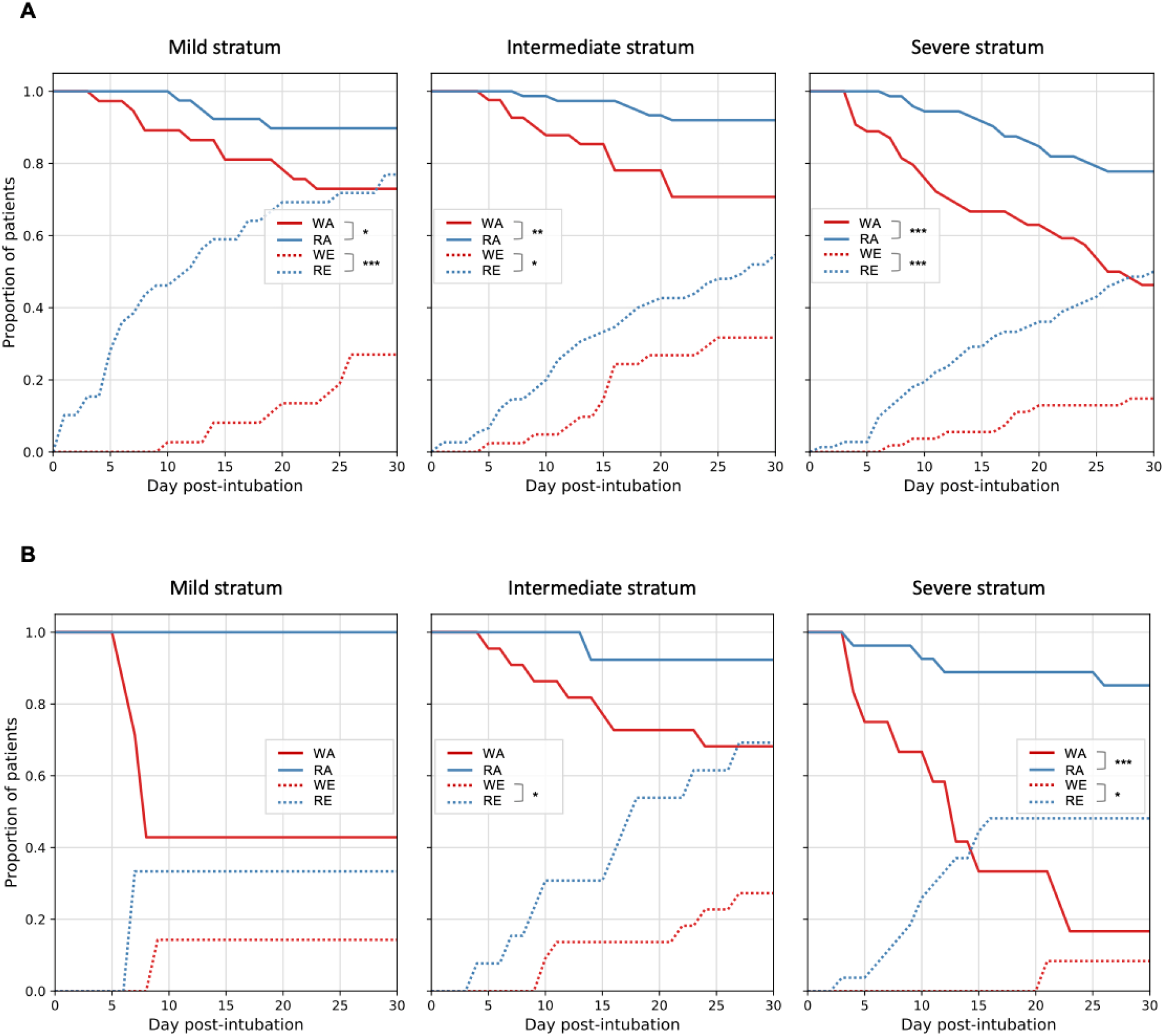
30-day outcomes (extubation, mortality, and tracheostomy) of the trajectory subphenotypes. Chi-square/Fisher’s exact tests were applied to compare 30-day outcomes between the worsening and recovering subphenotypes for each baseline strata. *(A)* 30-day outcomes of subphenotypes derived in NYP-WCMC cohort. *(B)* 30-day outcomes of subphenotypes derived in NYP-LMH validation cohort. * denoting testing significance passed p-value < 0.05; ** denoting testing significance passed p-value < 0.01; *** denoting testing significance passed p-value < 0.001. Abbreviations: WA=worsening subphenotype alive; RA=recovering subphenotype alive; WE=worsening subphenotype extubated; RE=recovering subphenotype extubated.

We assessed the differences between a broad range of laboratory tests, vital signs, and respiratory variables in the worsening and recovering subphenotypes. Importantly, basic laboratory tests and inflammatory markers were differentially associated with the worsening and recovering subphenotypes over time, which suggests that there is value in clinically following markers such as D-dimer, LDH, ferritin, procalcitonin and C-reactive protein. In the mild stratum, markers from the regular blood panel such as total white blood cell count and neutrophil counts, while inflammatory markers, such as ferritin and LDH, differentiate in the severe stratum. Laboratory tests, vitals and inflammatory markers in the intermediate stratum were less able to distinguish between the worsening and recovering subphenotypes. However, differences emerged over a longer time horizon (e.g., day 3). This further highlights the dynamic nature of COVID-19 and the difficulty in early prognosis in the critically ill population, despite severely deranged baseline organ dysfunction and inflammatory markers.

We built multivariable prediction models for the identified trajectory subphenotypes from patient baseline characteristics and early-stage clinical feature values. Models were built on at successive time points (day 1, 2, 3, 4, and 5) after intubation. Predictive performances measured by AUC-ROC improved as the number of days increased. The predictors’ importance to differentiating worsening and recovering subphenotypes showed varying patterns (Figure E-5). Importantly, aside from age and BMI, demographics, baseline comorbidities, and medications prescribed around intubation did not contribute to discriminating the subphenotypes in any of the strata. The persistence or development of renal failure, predicts subphenotype assignment later in the severe and intermediate strata, respectively. The persistence of vasodilatory shock in the intermediate stratum predicts the worsening subphenotype. While the development of thrombocytopenia, is discriminative late in the severe stratum. Interestingly, over the course of the first 7 days following intubation, liver failure remained rare. At different points in the course, inflammatory markers such as creatine kinase and D-dimer predicted worsening and recovering subphenotypes.

Our study was conducted on the two NYP system hospitals. Woresning and recovering SOFA subphenotypes, clinical characteristics, and outcomes from the validation cohort was consistent with the original subphenotypes. Although, due to the limited size of NYP-LMH validation cohort, statistical significance of some markers vanished, most of the results reflected the development cohort’s findings. This consistency ensures the existence of the worsening and recovering trajectory subphenotypes at each baseline stratum of the critically ill COVID-19 patients.

## Limitations

While this study presents a step forward in the efforts to parse the progression heterogeneity of critically ill patients with COVID-19, several limitations remain. The first limitation could be SOFA’s inadequacy in tabulating organ dysfunction in COVID-19 associated respiratory failure. For example, COVID-19 is associated with a different pattern of hypercoagulability compared to sepsis, which is reflected in the preserved platelet count in most this cohort’s patients.(33) Also despite elevations in liver chemistries in many patients, hyperbilirubinemia was rare. Despite this limitation, SOFA trajectory subphenotypes predicted mortality and will allow for future comparisons with other diseases.

Second, we did not use the progression of inflammatory markers such as C-reactive protein, D-dimer or ferritin, which are known risk factors for this disease, to identify the subphenotypes. Nor did we stratify patients based the severity of respiratory failure alone. Instead, we chose to see how these factors interacted with traditional organ dysfunction, as most patients with COVID19 die from multisystem organ failure and not refractory respiratory failure(8, 9).

Third, differentiating trajectory subphenotypes in this critically ill population was difficult, as AUC-ROC metrics of prediction modeling using data at day 1 post-intubation were around 0.7. By restricting our analysis to a very high-risk population, we decreased the discriminative power of many of our biomarkers to predict outcomes. All patients were high risk. However, we have added to our understanding of patients with critical COVID-19, by documenting the natural history of organ dysfunction in this population. Future research efforts, with novel biomarkers, are needed to predict worsening and recovering subphenotypes at an earlier time point in those with respiratory failure.

Fourth, the surge conditions in New York City during the study period could affect the study. Care may have been influenced by the surge conditions during this difficult time. However, all patients were cared for in a critical care environment and despite the massive patient burden, the all cause 30-day mortality was 25.9%.

## Conclusions

In a population of critically ill patients with COVID-19 respiratory failure, there are distinct worsening and recovering organ dysfunction trajectory subphenotypes. Worsening status was predictive of poor outcomes in all strata regardless of baseline severity. These findings highlight the importance of supportive care for sequential organ failure in addition to respiratory failure in this disease. Trajectory based subphenotypes offer a potential road map for understanding the evolution of critical illness in COVID-19. We call for further analysis.

## Data Availability

The patient data are from NYP-WCM and NYP-LMH and not publicly available.

## Acknowledgments

We thank the following Weill Cornell Medicine medical students for their contributions to the COVID-19 Registry through medical chart abstraction: Zara Adamou BA, Haneen Aljayyousi BA, Mark N. Alshak BA (student leader), Bryan K. Ang BA, Elena Beideck BS, Orrin S. Belden BS, Sharmi Biswas MD, Anthony F. Blackburn BS, Joshua W. Bliss PharmD, Kimberly A. Bogardus BA, Chelsea D. Boydstun BA, Clare A. Burchenal MPH, Eric T. Caliendo BS, John K. Chae BA, David L. Chang BS, Frank R. Chen BS, Kenny Chen BA, Andrew Cho PhD, Alice Chung BA, Alisha N. Dua MRes, Andrew Eidelberg BS, Rahmi S. Elahjji BA, Mahmoud Eljaby MMSc, Emily R. Eruysal BS, Kimberly N. Forlenza MSc, Rana Khan Fowlkes BA, Rachel L. Friedlander BA, Gary George BS, Shannon Glynn BS, Leora Haber BA, Janice Havasy BS, Alex Huang BA, Hao Huang BS, Jennifer H. Huang BS, Sonia Iosim BS, Mitali Kini BS, Rohini V. Kopparam BS, Jerry Y. Lee BA, Mark Lee BS BA, Aretina K. Leung BA, Han A. Li BA (student leader), Bethina Liu AB, Charalambia Louka BS, Brienne Lubor BS, Dianne Lumaquin BS, Matthew L. Magruder BA, Ruth Moges MSc, Prithvi M. Mohan BS, Max F. Morin BS, Sophie Mou BA, J. J. Nario BS, Yuna Oh BS, Noah Rossen BA, Emma M. Schatoff PhD, Pooja D. Shah BA, Sachin P. Shah BA, Daniel Skaf BS, Shoran Tamura BS, Ahmed Toure BA, Camila M. Villasante BA, Gal Wald BA, Graham T. Wehmeyer BS (student leader), Samuel Williams BA, Ashley Wu BS, Andrew L. Yin BA, Lisa Zhang BA

## Supplementary Materials

### Appendix 1. Patient care

Care of the patient was at the discretion of each attending physician. Patients were intubated prior to arrival to the ICU. Daily briefings were held during the surge to reinforce best practices and evidence related to caring for acute hypoxemic respiratory failure. Volume control ventilation targeting a tidal volume of 6-8 mls/kg IDBW and a plateau pressure < 30 cm/h20 was recommended. No particular PEEP titration protocol was mandated, however the ARDSnet moderate and high PEEP documents were made available to practitioners. Propofol was the first sedative of choice with the addition of opiates for dyspnea and perceived pain. An even to positive fluid balance was suggested on days one and two of mechanical ventilation. Patients with hypotension despite volume loading were treated with vasopressors, norepinephrine as the first choice to target a MAP of 65 mmhg. Prone positioning was suggested if a patient had a PF ratio of <150 despite optimization of PEEP and vent synchrony. Neuromuscular blockade was suggested in cases of refractory hypoxemia. Inhaled nitric oxide was available in cases of refractory respiratory failure at the discretion of the treating intensivists. Extra corporeal membrane oxygenation support was not offered due to resource limitations. HProphylactic anticoagulation was recommended for all patients and a higher dose prophylactic regimen was implemented during the course of the surge. Enoxaparin was the recommended first choice anticoagulation, even in the setting of renal insufficiency with strict monitoring of anti-factor Xa levels. Heparin was used for patients with concern for active bleeding or patients unable to tolerate enoxaparin. Corticosteroids were used at the discretion of the individual attending physician for the treatment of ARDS and for other traditional complications such as septic shock, bronchospasm and airway edema. Hydroxychloroquine without azithromycin was used to treat most patients as an antiviral. Remdesivir was available through compassionate use and through several clinical trials. Off label use of IL-6 inhibition was directed by consultation with infectious disease physicians. Choices regarding antibiotics and other therapeutics were at the discretion of treating intensivists with consultation with infectious disease physicians.

### Appendix 2. Data description

In this study, all data were collected from either the Weill Cornell-Critical carE Database for Advanced Research (WC-CEDAR)(1), Weill Cornell Medicine COVID Institutional Data Repository (COVID-IDR)(2), or via manual chart abstraction (REDCap). A wide range of data were included for analysis, including:

- Demographics include age, sex, race, and body mass index (BMI) were obtained when admitted to ICU.
- Chronic comorbidities were assessed at ICU admission and collected via chart abstraction. Comorbidities studied include coronary artery disease, cerebrovascular accident (stroke), heart failure, hypertension, diabetes mellitus, pulmonary disease, renal disease, cirrhosis, hepatitis, active cancer, transplant, inflammatory bowel disease, human immunodeficiency viruses, rheumatologic disease, and other immunosuppressed state.
- Medications prescribed were collected from the COVID-IDR database. Medications screened include tocilizumab, hydroxychloroquine, corticosteroids (such as prednisone, methylprednisolone, dexamethasone, and hydrocortisone), enoxaparin, heparin, and antibiotics (such as ceftriaxone, azithromycin, piperacillin-tazobactam, meropenem, vancomycin, and doxycycline). Enoxaparin was further assessed as prophylactic dose (<.5 mg per KG), high prophylactic dose (<1 mg per KG), and treatment dose (>1 mg per KG), while heparin was assessed as prophylactic dose (subcutaneous delivery) and treatment dose (intravenous delivery).
- In order to identify markers associated to identified subphenotypes, we assessed a broad spectrum of laboratory tests related to organ failure, including alanine aminotransferase (ALT), albumin level, aspartate aminotransferase (AST), total bilirubin, carbon dioxide, creatine kinase (CK) level, creatinine, C-reactive protein (CRP) level, D-dimer, erythrocyte sedimentation rate (ESR), ferritin level, globulin level, glucose level, hemoglobin level, lactate dehydrogenase (LDH), lactic acid level, lymphocyte percentage and count, neutrophil percentage and count, platelets level, potassium level, procalcitonin level, sodium level, troponin level, triglycerides, and white blood cell (WBC) count.
- Several vital signs were also assessed, including Glasgow Coma Scale score (GCS), and mean arterial pressure (MAP), temperature.
- Clinical variables reflecting respiratory status of patients were also studied, including fraction of inspired oxygen (FiO2), partial pressure of oxygen (PaO2), PaO2/FiO2 ratio (P/F ratio), and oxygen saturation (SpO2).
- Several parameters of ventilator setting were studied, including driving pressure, minute ventilation, arterial partial pressure of carbon dioxide (PCO2) level, positive end-expiratory pressure (PEEP), PH, peak inspiratory pressure (PIP), plateau pressure, static compliance, tidal volume, tidal volume to predicted body weight (PBW) ratio, and ventilator ratio.
- ABO/RH blood type data were collected from COVID-IDR database.

### Appendix 3. Inclusion exclusion criteria

#### Inclusion

We included patients with positive results on viral RNA detection by real-time reverse transcriptase polymerase chain reaction (RT-PCR) test from nasopharyngeal swabs specimens and treated with mechanical ventilation at the ICU in NYP-WCMC and NYP-LMH.

#### Exclusion

- Patients who were less than 18 years old when admitted to ICU were excluded for analysis.
- Since our aim was to identify organ dysfunction progression patterns within 7-days after intubation, missing too many (>50%) SOFA data may mislead understanding the trajectory trends. Hence, we excluded patients with ≤3 days SOFA data. Specifically, 20 patients of NYP-WCMC cohort were excluded, among which 9, 3, and 3 dead within day 1, 2, 3 after intubation, respectively, 2 have no records, and 2 and 1 only have 2- and 3-days SOFA data, respectively.
- 11 patients of NYP-LMH cohort were excluded, among which 2, 5, and 4 dead within day 1, 2, 3 after intubation, respectively.
- Outliers such as unchanged or heavily fluctuated trajectories which may hamper the model to capture clinically meaningful SOFA progression trends were excluded for analysis. Specifically, 7 unchanged trajectories of NYP-WCM cohort, and 3 and 5 heavily fluctuated trajectories within 7-day post-intubation of NYP-WCMC and NYP-LMH cohorts were excluded for analysis.

The overall inclusion exclusion criteria were provided in Figure E-1.

## Tables

**Table E-1.** Clustering metric (McClain index(3)) statistics and cluster number determination.

| # of clusters | NYP-WCMC cohort |  |  | NYP-LMH validation cohort |  |  |
| --- | --- | --- | --- | --- | --- | --- |
|  | Mild stratum | Intermediate stratum | Severe stratum | Mild stratum | Intermediate stratum | Severe stratum |
| 2 | 0.53 | 0.50 | 0.54 | 0.28 | 0.59 | 0.48 |
| 3 | 0.60 | 0.58 | 0.70 | 0.32 | 0.66 | 0.59 |
| 4 | 0.88 | 1.05 | 0.74 | 0.34 | 1.13 | 0.73 |
| 5 | 0.88 | 1.10 | 0.76 | 0.49 | 1.23 | 1.04 |
| 6 | 1.15 | 1.12 | 0.86 | 0.72 | 1.66 | 1.11 |
| **Optimal cluster #** | 2 | 2 | 2 | 2 | 2 | 2 |

The minimum value of the McClain index is used to indicate the optimal number of clusters.

**Table E-2.** Clinical characteristics of the trajectory subphenotypes in NYP-LMH validation cohort.

| Variable | Mild stratum<br>(SOFA 0-10, n=10) |  |  | Intermediate stratum<br>(SOFA 11-12, n=35) |  |  | Severe stratum<br>(SOFA 13-24, n=39) |  |  |
| --- | --- | --- | --- | --- | --- | --- | --- | --- | --- |
|  | Worsening | Recovering | p-value<sup>†</sup> | Worsening | Recovering | p-value<sup>†</sup> | Worsening | Recovering | p-value<sup>†</sup> |
| **Total #** | **7** | **3** | **-** | **22** | **13** | **-** | **12** | **27** | **-** |
| **Demographics** |  |  |  |  |  |  |  |  |  |
| Age, Mean (SD) | 60.29 (19.72) | 62.67 (7.93) | 0.861 | 60.73 (12.40) | 63.15 (9.47) | 0.559 | 78.00 (9.08) | 68.37 (10.58) | 0.011 |
| Sex female, n (%) | 4 (57.14%) | 0 (0.00%) | 0.200 | 10 (45.45%) | 9 (69.23%) | 0.293 | 2 (16.67%) | 8 (29.63%) | 0.693 |
| CAUCASIAN, n (%) | 0 (0.00%) | 0 (0.00%) | 1.000 | 2 (9.09%) | 2 (15.38%) | 0.149 | 2 (16.67%) | 1 (3.70%) | 0.345 |
| AFRICAN AMERICAN, n (%) | 0 (0.00%) | 0 (0.00%) |  | 0 (0.00%) | 0 (0.00%) |  | 3 (25.00%) | 4 (14.81%) |  |
| ASIAN / PACIFIC ISLANDER, n (%) | 3 (42.86%) | 2 (66.67%) |  | 9 (40.91%) | 3 (23.08%) |  | 3 (25.00%) | 12 (44.44%) |  |
| MULTI-RACIAL, n (%) | 2 (28.57%) | 0 (0.00%) |  | 1 (4.55%) | 4 (30.77%) |  | 0 (0.00%) | 3 (11.11%) |  |
| BMI, Mean (SD) | 26.77 (4.64) | 26.43 (1.14) | 0.914 | 31.22 (11.98) | 28.12 (4.57) | 0.262 | 26.90 (3.72) | 28.58 (6.25) | 0.409 |
| **Comorbidities** |  |  |  |  |  |  |  |  |  |
| Coronary Artery Disease, n (%) | 1 (14.29%) | 0 (0.00%) | 1.000 | 0 (0.00%) | 1 (7.69%) | 0.323 | 4 (33.33%) | 5 (18.52%) | 0.416 |
| Cerebrovascular accident (Stroke), n (%) | 0 (0.00%) | 0 (0.00%) | 1.000 | 0 (0.00%) | 0 (0.00%) | 1.000 | 3 (25.00%) | 1 (3.70%) | 0.078 |
| Heart Failure, n (%) | 0 (0.00%) | 0 (0.00%) | 1.000 | 0 (0.00%) | 1 (7.69%) | 0.323 | 0 (0.00%) | 2 (7.41%) | 1.000 |
| Hypertension, n (%) | 3 (42.86%) | 2 (66.67%) | 1.000 | 9 (40.91%) | 8 (61.54%) | 0.068 | 7 (58.33%) | 21 (77.78%) | 0.262 |
| Diabetes Mellitus, n (%) | 3 (42.86%) | 1 (33.33%) | 1.000 | 7 (31.82%) | 5 (38.46%) | 0.447 | 5 (41.67%) | 14 (51.85%) | 0.731 |
| Pulmonary Disease, n (%) | 1 (14.29%) | 1 (33.33%) | 1.000 | 2 (9.09%) | 2 (15.38%) | 0.577 | 3 (25.00%) | 6 (22.22%) | 1.000 |
| Renal Disease, n (%) | 0 (0.00%) | 0 (0.00%) | 1.000 | 0 (0.00%) | 2 (15.38%) | 0.097 | 0 (0.00%) | 5 (18.52%) | 0.299 |
| Cirrhosis, n (%) | 0 (0.00%) | 0 (0.00%) | 1.000 | 0 (0.00%) | 0 (0.00%) | 1.000 | 0 (0.00%) | 1 (3.70%) | 1.000 |
| Hepatitis, n (%) | 0 (0.00%) | 0 (0.00%) | 1.000 | 1 (4.55%) | 0 (0.00%) | 1.000 | 0 (0.00%) | 1 (3.70%) | 1.000 |
| HIV, n (%) | 0 (0.00%) | 0 (0.00%) | 1.000 | 1 (4.55%) | 0 (0.00%) | 1.000 | 0 (0.00%) | 0 (0.00%) | 1.000 |
| Active Cancer, n (%) | 0 (0.00%) | 0 (0.00%) | 1.000 | 0 (0.00%) | 0 (0.00%) | 1.000 | 1 (8.33%) | 1 (3.70%) | 0.526 |
| Transplant, n (%) | 0 (0.00%) | 0 (0.00%) | 1.000 | 0 (0.00%) | 0 (0.00%) | 1.000 | 0 (0.00%) | 1 (3.70%) | 1.000 |
| Inflammatory Bowel Disease, n (%) | 0 (0.00%) | 0 (0.00%) | 1.000 | 0 (0.00%) | 0 (0.00%) | 1.000 | 0 (0.00%) | 0 (0.00%) | 1.000 |
| Rheumatologic Disease, n (%) | 0 (0.00%) | 0 (0.00%) | 1.000 | 0 (0.00%) | 2 (15.38%) | 0.097 | 1 (8.33%) | 0 (0.00%) | 0.308 |
| Other Immunosuppressed State, n (%) | 0 (0.00%) | 0 (0.00%) | 1.000 | 0 (0.00%) | 0 (0.00%) | 1.000 | 0 (0.00%) | 0 (0.00%) | 1.000 |
| **Baseline SOFA scores** |  |  |  |  |  |  |  |  |  |
| Cardiovascular, Mean (SD) | 1.43 (1.05) | 1.33 (0.94) | 0.500 | 3.50 (0.78) | 3.69 (0.82) | 0.169 | 4.00 (0.00) | 3.81 (0.47) | 0.087 |
| Central nervous system, Mean (SD) | 2.29 (1.48) | 3.33 (0.47) | 0.183 | 3.27 (0.45) | 3.54 (0.50) | 0.063 | 3.42 (0.49) | 3.70 (0.53) | 0.039 |
| Coagulation, Mean (SD) | 0.00 (0.00) | 0.00 (0.00) | - | 0.18 (0.49) | 0.00 (0.00) | 0.091 | 0.33 (0.62) | 0.11 (0.31) | 0.127 |
| Liver, Mean (SD) | 0.14 (0.35) | 0.00 (0.00) | 0.331 | 0.23 (0.52) | 0.00 (0.00) | 0.057 | 0.42 (0.64) | 0.22 (0.50) | 0.160 |
| Renal, Mean (SD) | 0.43 (0.73) | 0.67 (0.47) | 0.257 | 0.36 (0.48) | 0.38 (0.84) | 0.270 | 2.33 (0.85) | 2.52 (1.17) | 0.307 |
| Respiration, Mean (SD) | 4.00 (0.00) | 4.00 (0.00) | - | 3.91 (0.42) | 4.00 (0.00) | 0.239 | 4.00 (0.00) | 4.00 (0.00) | - |
| SOFA score, Mean (SD) | 8.29 (2.43) | 9.33 (0.47) | 0.500 | 11.45 (0.50) | 11.62 (0.49) | 0.188 | 14.50 (1.04) | 14.37 (1.09) | 0.377 |

<sup>†</sup> p-value calculated by Chi-square test/Fisher's exact test, or student's t-test/Mann-Whitney test where appropriate.
<sup>\*\*</sup> False discovery rate corrected p-value < 0.05
Abbreviation: BMI=body mass index, HIV=Human Immunodeficiency Virus, NYP-LMH=New York Presbyterian Hospital-Lower Manhattan Hospital, SD=standard deviation, SOFA=Sequential Organ Failure Assessment

**Table E-3.** Medication of the trajectory subphenotypes in NYP-WCMC cohort.

| Medication | Mild stratum<br>(SOFA 0-10, n=76) |  |  | Intermediate stratum<br>(SOFA 11-12, n=116) |  |  | Severe stratum<br>(SOFA 13-24, n=126) |  |  |
| --- | --- | --- | --- | --- | --- | --- | --- | --- | --- |
|  | Worsening<br>(n=39) | Recovering<br>(n=37) | p-value<sup>†</sup> | Worsening<br>(n=41) | Recovering<br>(n=75) | p-value<sup>†</sup> | Worsening<br>(n=54) | Recovering<br>(n=72) | p-value<sup>†</sup> |
| Tocilizumab, n (%) | 4 (10.81%) | 3 (7.69%) | 0.708 | 5 (12.20%) | 5 (6.67%) | 0.321 | 4 (7.41%) | 4 (5.56%) | 0.724 |
| Hydroxychloroquine, n (%) | 29 (78.38%) | 27 (69.23%) | 0.439 | 35 (85.37%) | 51 (68.00%) | 0.048 | 41 (75.93%) | 52 (72.22%) | 0.686 |
| Prednisone, n (%) | 0 (0.00%) | 4 (10.26%) | 0.116 | 1 (2.44%) | 1 (1.33%) | 1.000 | 3 (5.56%) | 2 (2.78%) | 0.651 |
| Methylprednisolone, n (%) | 7 (18.92%) | 10 (25.64%) | 0.586 | 8 (19.51%) | 16 (21.33%) | 1.000 | 11 (20.37%) | 8 (11.11%) | 0.208 |
| Dexamethasone, n (%) | 1 (2.70%) | 2 (5.13%) | 1.000 | 1 (2.44%) | 4 (5.33%) | 0.655 | 0 (0.00%) | 2 (2.78%) | 0.506 |
| Hydrocortisone, n (%) | 5 (13.51%) | 1 (2.56%) | 0.103 | 7 (17.07%) | 6 (8.00%) | 0.216 | 15 (27.78%) | 8 (11.11%) | 0.020 |
| Ceftriaxone, n (%) | 18 (48.65%) | 21 (53.85%) | 0.819 | 23 (56.10%) | 43 (57.33%) | 1.000 | 25 (46.30%) | 40 (55.56%) | 0.369 |
| Azithromycin, n (%) | 8 (21.62%) | 7 (17.95%) | 0.777 | 5 (12.20%) | 13 (17.33%) | 0.595 | 8 (14.81%) | 17 (23.61%) | 0.264 |
| Piperacillin-tazobactam, n (%) | 12 (32.43%) | 13 (33.33%) | 1.000 | 25 (60.98%) | 29 (38.67%) | 0.032 | 24 (44.44%) | 36 (50.00%) | 0.591 |
| Meropenem, n (%) | 4 (10.81%) | 4 (10.26%) | 1.000 | 3 (7.32%) | 3 (4.00%) | 0.664 | 12 (22.22%) | 7 (9.72%) | 0.077 |
| Vancomycin, n (%) | 10 (27.03%) | 12 (30.77%) | 0.803 | 25 (60.98%) | 23 (30.67%) | 0.003 | 24 (44.44%) | 32 (44.44%) | 1.000 |
| Doxycycline, n (%) | 19 (51.35%) | 22 (56.41%) | 0.818 | 19 (46.34%) | 34 (45.33%) | 1.000 | 21 (38.89%) | 34 (47.22%) | 0.370 |
| Enoxaparin, n (%) |  |  |  |  |  |  |  |  |  |
| Prophylactic dose (<.5 mg/Kg) | 8 (21.62%) | 10 (25.64%) |  | 9 (21.95%) | 15 (20.00%) |  | 14 (25.93%) | 17 (23.61%) |  |
| High prophylactic dose (>.5 mg/Kg) | 13 (35.14%) | 21 (53.85%) | 0.117 | 17 (41.46%) | 37 (49.33%) | 0.824 | 10 (18.52%) | 28 (38.89%) | 0.241 |
| Treatment dose (>1 mg/Kg) | 8 (21.62%) | 6 (15.38%) |  | 8 (19.51%) | 14 (18.67%) |  | 7 (12.96%) | 7 (9.72%) |  |
| Heparin, n (%) |  |  |  |  |  |  |  |  |  |
| Prophylactic dose (subcutaneous) | 5 (13.51%) | 9 (23.08%) |  | 13 (31.71%) | 9 (12.00%) |  | 18 (33.33%) | 25 (34.72%) |  |
| treatment dose | 3 (8.11%) | 3 (7.69%) | 0.539 | 8 (19.51%) | 2 (2.67%) | <0.001** | 18 (33.33%) | 14 (19.44%) | 0.170 |

Medications were prescribed within the window from 3-day before to 5-day after intubation.
<sup>†</sup> p-value calculated by Chi-square/Fisher's exact test
\*\* False discovery rate corrected p-value < 0.05
Abbreviation: BMI=body mass index, HIV=Human Immunodeficiency Virus, NYP-WCMC=New York Presbyterian Hospital-Weill Cornell Medical Center, SD=standard deviation, SOFA=Sequential Organ Failure Assessment

**Table E-4.** Medication of the trajectory subphenotypes in NYP-LMH cohort.

| Medication | Mild stratum<br>(SOFA 0-10, n=10) |  |  | Intermediate stratum<br>(SOFA 11-12, n=35) |  |  | Severe stratum<br>(SOFA 13-24, n=39) |  |  |
| --- | --- | --- | --- | --- | --- | --- | --- | --- | --- |
|  | Worsening<br>(n=7) | Recovering<br>(n=3) | p-value<sup>†</sup> | Worsening<br>(n=22) | Recovering<br>(n=13) | p-value<sup>†</sup> | Worsening<br>(n=12) | Recovering<br>(n=27) | p-value<sup>†</sup> |
| Tocilizumab, n (%) | 0 (0.00%) | 0 (0.00%) | 1.000 | 4 (18.18%) | 1 (7.69%) | 0.635 | 1 (8.33%) | 5 (18.52%) | 0.645 |
| Hydroxychloroquine, n (%) | 6 (85.71%) | 3 (100.00%) | 1.000 | 19 (86.36%) | 11 (84.62%) | 1.000 | 10 (83.33%) | 22 (81.48%) | 1.000 |
| Prednisone, n (%) | 0 (0.00%) | 0 (0.00%) | 1.000 | 0 (0.00%) | 0 (0.00%) | 1.000 | 0 (0.00%) | 1 (3.70%) | 1.000 |
| Methylprednisolone, n (%) | 2 (28.57%) | 2 (66.67%) | 0.500 | 8 (36.36%) | 3 (23.08%) | 0.705 | 2 (16.67%) | 7 (25.93%) | 0.693 |
| Dexamethasone, n (%) | 0 (0.00%) | 1 (33.33%) | 0.300 | 0 (0.00%) | 0 (0.00%) | 1.000 | 0 (0.00%) | 0 (0.00%) | 1.000 |
| Hydrocortisone, n (%) | 1 (14.29%) | 0 (0.00%) | 1.000 | 0 (0.00%) | 1 (7.69%) | 0.353 | 5 (41.67%) | 6 (22.22%) | 0.262 |
| Ceftriaxone, n (%) | 3 (42.86%) | 3 (100.00%) | 0.200 | 17 (77.27%) | 9 (69.23%) | 1.000 | 6 (50.00%) | 15 (55.56%) | 1.000 |
| Azithromycin, n (%) | 4 (57.14%) | 1 (33.33%) | 1.000 | 11 (50.00%) | 4 (30.77%) | 0.476 | 4 (33.33%) | 8 (29.63%) | 1.000 |
| Piperacillin-tazobactam, n (%) | 5 (71.43%) | 1 (33.33%) | 0.500 | 11 (50.00%) | 8 (61.54%) | 0.476 | 8 (66.67%) | 14 (51.85%) | 0.494 |
| Meropenem, n (%) | 0 (0.00%) | 0 (0.00%) | 1.000 | 0 (0.00%) | 1 (7.69%) | 0.353 | 0 (0.00%) | 0 (0.00%) | 1.000 |
| Vancomycin, n (%) | 3 (42.86%) | 1 (33.33%) | 1.000 | 9 (40.91%) | 8 (61.54%) | 0.282 | 10 (83.33%) | 15 (55.56%) | 0.151 |
| Doxycycline, n (%) | 2 (28.57%) | 2 (66.67%) | 0.500 | 10 (45.45%) | 6 (46.15%) | 1.000 | 7 (58.33%) | 11 (40.74%) | 0.488 |
| Enoxaparin, n (%) |  |  |  |  |  |  |  |  |  |
| Prophylactic dose (<.5 mg/Kg) | 2 (28.57%) | 0 (0.00%) |  | 10 (45.45%) | 5 (38.46%) |  | 3 (25.00%) | 8 (29.63%) |  |
| High prophylactic dose (>.5 mg/Kg) | 4 (57.14%) | 2 (66.67%) | 1.000 | 8 (36.36%) | 7 (53.85%) | 0.446 | 1 (8.33%) | 10 (37.04%) | 0.075 |
| Treatment dose (>1 mg/Kg) | 0 (0.00%) | 0 (0.00%) |  | 1 (4.55%) | 0 (0.00%) |  | 1 (8.33%) | 4 (14.81%) |  |
| Heparin, n (%) |  |  |  |  |  |  |  |  |  |
| Prophylactic dose (subcutaneous) | 4 (57.14%) | 1 (33.33%) | 1.000 | 12 (54.55%) | 5 (38.46%) | 0.205 | 7 (58.33%) | 11 (40.74%) | 0.503 |
| treatment dose | 0 (0.00%) | 0 (0.00%) |  | 3 (13.64%) | 0 (0.00%) |  | 3 (25.00%) | 7 (25.93%) |  |

Medications were prescribed within the window from 3-day before to 5-day after intubation.
<sup>†</sup> p-value calculated by Chi-square/Fisher's exact test
\*\* False discovery rate corrected p-value < 0.05
Abbreviation: BMI=body mass index, HIV=Human Immunodeficiency Virus, NYP-LMH=New York Presbyterian Hospital-Lower Manhattan Hospital, SD=standard deviation, SOFA=Sequential Organ Failure Assessment

**Table E-5.** Clinical variables (laboratory test results, vital signs, respiratory variables) of the baseline strata in NYP-WCMC cohort. Data were examined at day 1 post-intubation.

| Variable | All | Mild stratum | Intermediate stratum | Severe stratum | p-value<sup>†</sup> | Post-hoc |
| --- | --- | --- | --- | --- | --- | --- |
| ALT, IU/L, Median [IQR] | 42.0 [26.0-70.75] | 39.0 [23.75-59.12] | 42.0 [27.38-70.25] | 44.5 [26.25-74.0] | 0.631 |  |
| Albumin, g/dL, Median [IQR] | 2.1 [1.8-2.49] | 2.2 [1.9-2.5] | 2.2 [1.94-2.4] | 2.0 [1.7-2.45] | 0.109 |  |
| AST, IU/L, Median [IQR] | 49.0 [35.0-79.5] | 40.5 [33.5-61.0] | 48.0 [34.75-65.75] | 59.75 [35.0-98.0] | 0.005<sup>**,\$</sup> | Severe vs Mild, Intermediate |
| Bilirubin, mg/dL, Median [IQR] | 0.7 [0.5-1.0] | 0.6 [0.5-0.9] | 0.7 [0.47-0.9] | 0.78 [0.5-1.2] | 0.247 |  |
| Carbon dioxide, mmol/L, Median [IQR] | 25.42 [23.0-28.0] | 26.75 [25.0-29.0] | 26.0 [23.25-28.62] | 24.0 [21.75-26.38] | <0.001<sup>**,\$</sup> | Severe vs Mild, Intermediate |
| CK, U/L, Median [IQR] | 181.0 [77.0-326.75] | 172.25 [61.75-253.5] | 154.25 [79.75-274.63] | 187.0 [77.25-407.75] | 0.177 |  |
| Creatinine, mg/dL, Median [IQR] | 1.1 [0.8-1.77] | 0.81 [0.67-0.97] | 0.98 [0.77-1.26] | 1.86 [1.29-3.43] | <0.001<sup>**,\$</sup> | All comparison |
| CRP, mg/dL, Median [IQR] | 20.25 [12.95-27.9] | 18.65 [10.8-26.23] | 20.94 [13.57-27.52] | 20.11 [13.6-28.08] | 0.428 |  |
| D-dimer, ng/mL, Median [IQR] | 1331.75 [734.0-3185.0] | 1087.5 [596.75-2373.95] | 1331.75 [692.38-3023.5] | 1942.62 [1009.0-4408.14] | 0.003<sup>**,\$</sup> | Severe vs Mild, Intermediate |
| ESR, mm/hr, Median [IQR] | 83.0 [67.25-97.38] | 83.0 [72.92-100.25] | 83.0 [67.0-89.06] | 83.0 [66.58-100.0] | 0.243 |  |
| Ferritin, ng/mL, Median [IQR] | 1166.3 [772.52-1639.7] | 1121.9 [601.9-1611.3] | 1166.3 [636.66-1596.6] | 1283.45 [927.74-1766.8] | 0.014<sup>**,\$</sup> | Severe vs Mild, Intermediate |
| Globulin, g/dL, Median [IQR] | 3.4 [3.1-3.8] | 3.3 [3.1-3.7] | 3.4 [3.14-3.8] | 3.4 [3.0-3.89] | 0.503 |  |
| Glucose, mg/dL, Median [IQR] | 142.5 [113.5-190.38] | 118.25 [103.88-146.25] | 145.5 [119.25-189.25] | 154.0 [124.0-207.5] | <0.001<sup>**,\$</sup> | Mild vs Intermediate, Severe |
| Hemoglobin, g/dL, Median [IQR] | 11.8 [10.22-12.9] | 11.82 [10.44-12.91] | 11.72 [10.57-12.9] | 11.68 [9.5-12.7] | 0.294 |  |
| LDH, U/L, Median [IQR] | 500.0 [432.0-667.25] | 472.5 [388.0-556.29] | 498.5 [439.19-649.58] | 560.75 [460.25-788.0] | 0.001<sup>**,\$</sup> | Mild vs Intermediate, Severe |
| Lactic acid level, mmol/L, Mean [CI] | 1.6 [1.52,1.68] | 1.61 [1.48,1.75] | 1.5 [1.38,1.63] | 1.67 [1.53,1.82] | 0.197 |  |
| Lymphocyte percentage, %, Median [IQR] | 7.58 [4.53-12.1] | 7.3 [4.65-11.9] | 8.37 [5.28-12.8] | 6.95 [3.91-11.88] | 0.367 |  |
| Lymphocyte count, x10(9)/L, Median [IQR] | 0.8 [0.5-1.18] | 0.68 [0.49-1.09] | 0.9 [0.55-1.22] | 0.75 [0.49-1.17] | 0.098 |  |
| Neutrophil percentage, %, Median [IQR] | 86.0 [80.0-89.86] | 85.8 [78.6-89.77] | 85.25 [79.68-89.26] | 86.97 [80.4-90.69] | 0.301 |  |
| Neutrophil count, x10(9)/L, Median [IQR] | 9.19 [6.79-12.43] | 7.91 [6.21-10.65] | 9.11 [7.15-12.49] | 9.78 [7.12-13.84] | 0.017<sup>**,\$</sup> | Mild vs Intermediate, Severe |
| Platelet, x10(9)/L, Median [IQR] | 249.5 [182.25-334.5] | 251.5 [193.5-326.75] | 266.0 [211.75-363.75] | 230.0 [169.12-300.75] | 0.003<sup>**,\$</sup> | Intermediate vs Severe |
| Potassium, mmol/L, Median [IQR] | 4.27 [3.9-4.65] | 4.18 [3.8-4.5] | 4.26 [3.89-4.54] | 4.4 [3.9-4.93] | 0.009<sup>**,\$</sup> | Severe vs Mild, Intermediate |
| Procalcitonin, ng/mL, Median [IQR] | 0.74 [0.31-2.25] | 0.43 [0.17-0.9] | 0.52 [0.27-1.05] | 1.92 [0.63-7.01] | <0.001<sup>**§</sup> | Severe vs Mild, Intermediate |
| Sodium, mmol/L, Median [IQR] | 140.0 [136.27-142.5] | 140.58 [136.88-142.62] | 139.33 [137.0-142.0] | 139.5 [136.0-143.0] | 0.402 |  |
| Troponin, ng/mL, Mean [CI] | 1.45 [0.52,2.37] | 0.38 [0.08,0.68] | 0.46 [0.08,0.84] | 3.0 [0.71,5.29] | 0.001<sup>**§</sup> | Mild vs Intermediate, Severe |
| Triglycerides, mg/dL, Median [IQR] | 224.5 [156.12-296.75] | 237.0 [147.5-291.0] | 218.12 [161.5-296.75] | 224.5 [162.5-300.25] | 0.997 |  |
| White blood cell, x10(9)/L, Median [IQR] | 11.18 [8.1-14.9] | 10.4 [7.07-12.49] | 11.07 [8.45-14.95] | 12.12 [8.96-15.8] | 0.009<sup>**</sup> | Mild vs Intermediate, Severe |
| GCS, Mean [CI] | 4.04 [3.79,4.29] | 5.2 [4.4,6.0] | 4.14 [3.78,4.5] | 3.25 [3.1,3.41] | <0.001<sup>**§</sup> | Severe vs Mild, Intermediate |
| MAP, mmHg, Median [IQR] | 82.26 [76.62-88.13] | 83.9 [78.21-91.99] | 82.6 [77.19-87.06] | 80.89 [75.5-87.74] | 0.028<sup>§</sup> | Mild vs Severe |
| Temperature, degrees C, Median [IQR] | 37.04 [36.58-37.66] | 37.06 [36.6-37.76] | 37.08 [36.7-37.68] | 36.97 [36.53-37.56] | 0.314 |  |
| Urine output, mL, Median [IQR] | 1115.0 [741.25-1615.0] | 1210.0 [942.5-1761.25] | 1332.5 [921.25-1703.75] | 840.5 [391.75-1264.75] | <0.001<sup>**§</sup> | Severe vs Mild, Intermediate |
| FiO2, %, Median [IQR] | 64.65 [53.59-74.47] | 63.74 [51.09-74.34] | 66.12 [56.01-73.11] | 64.74 [54.41-75.17] | 0.693 |  |
| PaO2, mmHg, Median [IQR] | 89.0 [76.67-108.83] | 89.08 [77.0-112.31] | 86.28 [76.47-105.12] | 91.42 [75.99-108.83] | 0.733 |  |
| P/F ratio, Median [IQR] | 144.02 [114.24-183.91] | 152.0 [122.08-200.9] | 137.81 [114.55-178.16] | 142.89 [111.56-183.35] | 0.406 |  |
| SpO2, Median [IQR] | 96.1 [94.44-97.44] | 96.63 [94.53-97.69] | 95.78 [94.32-97.28] | 95.75 [94.49-97.28] | 0.280 |  |
| Driving pressure, Median [IQR] | 13.17 [11.0-15.86] | 13.08 [9.73-15.04] | 13.9 [11.9-16.54] | 13.17 [11.54-15.19] | 0.038 | Mild vs Intermediate |
| Minute ventilation, L, Median [IQR] | 9.98 [8.6-11.3] | 9.95 [8.69-10.46] | 9.77 [8.52-11.12] | 10.09 [8.75-11.83] | 0.178 |  |
| PCO2, mmHg, Median [IQR] | 45.92 [40.75-52.38] | 44.9 [39.88-48.31] | 46.14 [41.0-53.12] | 46.86 [40.6-53.0] | 0.194 |  |
| PEEP, Median [IQR] | 10.83 [9.45-12.73] | 10.0 [8.6-12.12] | 11.0 [10.0-12.88] | 11.1 [9.33-12.73] | 0.213 |  |
| PH, Mean [CI] | 7.37 [7.36,7.37] | 7.37 [7.37,7.38] | 7.37 [7.37,7.38] | 7.36 [7.35,7.37] | 0.029<sup>§</sup> | Severe vs Mild, Intermediate |
| PIP, Median [IQR] | 30.0 [26.0-33.33] | 29.33 [23.92-32.5] | 29.83 [26.0-34.21] | 30.37 [26.89-33.3] | 0.017<sup>§</sup> | Mild vs Severe |
| Plateau pressure, Median [IQR] | 25.0 [22.0-28.0] | 24.0 [20.38-27.0] | 25.5 [23.0-28.0] | 25.0 [22.0-28.0] | 0.066 |  |
| Static compliance, Median [IQR] | 29.67 [23.22-37.02] | 31.47 [24.59-43.55] | 28.57 [23.07-35.0] | 29.71 [21.7-37.31] | 0.083 |  |
| Tidal PBW ratio, Median [IQR] | 6.68 [6.34-7.52] | 6.68 [6.48-7.52] | 6.76 [6.35-7.7] | 6.68 [6.29-7.3] | 0.439 |  |
| Tidal volume, mL, Median [IQR] | 433.67 [397.66-473.75] | 437.92 [405.62-465.0] | 425.36 [380.0-468.12] | 445.5 [390.62-477.25] | 0.608 |  |
| Ventilator ratio, Median [IQR] | 2.04 [1.64-2.36] | 1.98 [1.58-2.1] | 1.93 [1.58-2.35] | 2.1 [1.69-2.45] | 0.144 |  |

<sup>†</sup> p-value calculated by analysis of variance (ANOVA)/Kruskal–Wallis test
<sup>\*\*</sup> False discovery rate corrected p-value < 0.05
<sup>§</sup> Age adjusted (analysis of covariance [ANCOVA]) p-value < 0.05
Abbreviations: ALT=Alanine aminotransferase, AST=Aspartate aminotransferase, CI=confidence interval, CK=Creatine kinase, CRP=C-reactive protein, ESR=Erythrocyte sedimentation rate, FiO2=fraction of inspired oxygen, GCS=Glasgow Coma Scale, IQR=Interquartile range, LDH=Lactate dehydrogenase, MAP=Mean arterial pressure,
PaO<sub>2</sub>=partial pressure of oxygen, PBW=predicted body weight, PCO<sub>2</sub>=arterial partial pressure of carbon dioxide, PEEP=Positive end-expiratory pressure, PIP=Peak inspiratory pressure, P/F ratio=PaO<sub>2</sub>/FiO<sub>2</sub> ratio, SpO<sub>2</sub>=oxygen saturation.

**Table E-6.** Clinical variables (laboratory test results, vital signs, respiratory variables) of the baseline strata in NYP-LMH cohort. Data were examined at day 1 post-intubation.

| Variable | All | Mild stratum | Intermediate stratum | Severe stratum | p-value<sup>†</sup> | Post-hoc |
| --- | --- | --- | --- | --- | --- | --- |
| ALT, IU/L, Median [IQR] | 38.0 [24.0-61.25] | 64.5 [49.0-77.5] | 33.0 [21.75-49.17] | 40.0 [24.0-61.25] | 0.045 | Mild vs Intermediate |
| Albumin, g/dL, Median [IQR] | 3.1 [2.72-3.3] | 3.1 [3.06-3.24] | 3.1 [2.7-3.47] | 3.0 [2.7-3.2] | 0.211 |  |
| AST, IU/L, Median [IQR] | 54.0 [39.25-79.0] | 58.0 [47.5-68.94] | 53.0 [36.5-73.0] | 51.33 [37.0-93.67] | 0.782 |  |
| Bilirubin, mg/dL, Median [IQR] | 0.7 [0.5-0.9] | 0.7 [0.55-0.88] | 0.6 [0.5-0.8] | 0.65 [0.45-1.1] | 0.873 |  |
| Carbon dioxide, mmol/L, Median [IQR] | 20.42 [18.5-23.0] | 20.5 [20.0-21.62] | 22.0 [19.75-24.5] | 19.8 [16.25-22.0] | 0.016 | Intermediate vs Severe |
| CK, U/L, Median [IQR] | 210.0 [101.65-481.81] | 287.5 [140.5-1026.75] | 240.0 [101.1-426.83] | 184.0 [110.17-466.0] | 0.690 |  |
| Creatinine, mg/dL, Median [IQR] | 1.4 [0.8-2.23] | 1.0 [0.7-1.25] | 0.9 [0.7-1.25] | 2.3 [1.6-2.85] | <0.001<sup>**,§</sup> | Severe vs Mild, Intermediate |
| CRP, mg/dL, Median [IQR] | 17.87 [15.12-24.52] | 16.27 [15.5-17.55] | 18.19 [14.68-25.12] | 18.61 [15.05-24.14] | 0.661 |  |
| D-dimer, ng/mL, Median [IQR] | 1508.0 [979.38-2955.53] | 1329.0 [679.38-1508.0] | 1470.0 [710.0-2457.54] | 1747.0 [1207.25-3857.5] | 0.053 |  |
| ESR, mm/hr, Median [IQR] | 88.0 [69.88-98.92] | 88.0 [79.75-88.0] | 88.0 [73.0-97.5] | 88.0 [64.0-109.0] | 0.954 |  |
| Ferritin, ng/mL, Median [IQR] | 1242.57 [889.56-1901.14] | 1330.89 [1099.24-2126.9] | 1426.04 [995.66-1935.52] | 1185.5 [702.08-1595.1] | 0.430 |  |
| Globulin, g/dL, Median [IQR] | 3.0 [3.0-3.5] | 3.0 [2.5-3.0] | 3.0 [3.0-3.5] | 3.0 [3.0-3.5] | 0.331 |  |
| Glucose, mg/dL, Median [IQR] | 150.5 [114.88-196.62] | 150.0 [123.31-188.75] | 121.0 [100.5-171.25] | 172.0 [131.33-228.95] | 0.005<sup>§</sup> | Intermediate vs Severe |
| Hemoglobin, g/dL, Median [IQR] | 12.05 [11.1-13.25] | 12.62 [12.03-13.03] | 11.9 [11.18-12.95] | 12.27 [10.8-13.62] | 0.684 |  |
| LDH, U/L, Median [IQR] | 518.39 [437.12-741.75] | 546.71 [485.88-678.25] | 504.0 [430.42-673.5] | 561.5 [437.17-784.75] | 0.719 |  |
| Lactic acid level, mmol/L, Median [IQR] | 2.2 [1.88,2.52] | 2.95 [1.29,4.62] | 2.01 [1.55,2.47] | 2.17 [1.75,2.6] | 0.312 |  |
| Lymphocyte percentage, %, Median [IQR] | 8.15 [4.64-13.29] | 7.55 [5.12-11.67] | 9.55 [7.33-15.0] | 5.9 [3.92-11.47] | 0.008 | Intermediate vs Severe |
| Lymphocyte count, x10(9)/L, Median [IQR] | 0.74 [0.53-1.06] | 0.7 [0.6-0.88] | 0.81 [0.63-1.2] | 0.7 [0.46-1.0] | 0.225 |  |
| Neutrophil percentage, %, Median [IQR] | 86.88 [80.45-91.14] | 87.65 [82.46-90.7] | 85.4 [79.6-87.38] | 90.05 [83.0-92.05] | 0.007 | Intermediate vs Severe |
| Neutrophil count, x10(9)/L, Median [IQR] | 8.88 [6.27-11.34] | 8.76 [6.54-12.22] | 7.75 [5.59-9.61] | 10.37 [6.73-12.81] | 0.013<sup>§</sup> | Intermediate vs Severe |
| Platelet, x10(9)/L, Median [IQR] | 236.0 [187.75-312.25] | 202.5 [188.25-292.5] | 264.0 [188.0-317.5] | 221.0 [177.5-303.0] | 0.463 |  |
| Potassium, mmol/L, Median [IQR] | 4.2 [3.8-4.67] | 4.38 [3.95-4.66] | 4.0 [3.7-4.35] | 4.23 [3.92-5.05] | 0.016<sup>§</sup> | Intermediate vs Severe |
| Procalcitonin, ng/mL, Median [IQR] | 0.8 [0.3-1.82] | 0.36 [0.15-0.57] | 0.49 [0.17-2.01] | 1.11 [0.46-1.84] | 0.038 | Mild vs Severe |
| Sodium, mmol/L, Median [IQR] | 138.25 [135.0-141.54] | 139.88 [136.25-142.75] | 139.0 [134.0-141.0] | 138.0 [135.5-140.75] | 0.693 |  |
| Troponin, ng/mL, Mean [CI] | 0.33 [0.13,0.54] | 0.04 [0.0,0.08] | 0.31 [0.02,0.61] | 0.43 [0.06,0.8] | 0.071 |  |
| Triglycerides, mg/dL, Median [IQR] | 228.0 [157.25-311.25] | 229.0 [228.0-310.14] | 228.0 [160.0-301.0] | 228.0 [152.0-309.0] | 0.566 |  |
| White blood cell, x10(9)/L, Median [IQR] | 10.8 [7.79-13.56] | 8.95 [6.9-13.43] | 10.4 [7.45-11.9] | 12.4 [8.6-14.45] | 0.081 |  |
| GCS, Mean [CI] | 5.36 [4.81,5.91] | 7.8 [4.83,10.77] | 5.51 [4.87,6.16] | 4.59 [3.9,5.28] | 0.010<sup>§</sup> | Severe vs Mild, Intermediate |
| MAP, mmHg, Median [IQR] | 82.91 [77.78-86.68] | 83.96 [78.58-91.71] | 82.52 [76.82-85.95] | 84.8 [79.18-88.49] | 0.238 |  |
| Temperature, degrees C, Median [IQR] | 37.19 [36.7-37.64] | 37.13 [36.46-37.62] | 37.42 [36.95-37.73] | 37.03 [36.63-37.43] | 0.121 |  |
| Urine output, mL, Median [IQR] | 947.5 [601.25-1400.0] | 1227.5 [1119.38-1612.5] | 1120.0 [882.5-1555.0] | 620.0 [310.0-952.5] | <0.001<sup>**§</sup> | Severe vs Mild, Intermediate |
| FiO2, %, Median [IQR] | 77.88 [66.67-88.87] | 68.74 [59.2-87.44] | 75.0 [62.0-89.38] | 80.0 [71.88-89.75] | 0.157 |  |
| PaO2, mmHg, Median [IQR] | 111.04 [89.5-134.83] | 105.69 [96.09-129.74] | 98.62 [86.5-122.92] | 118.0 [95.58-138.09] | 0.238 |  |
| P/F ratio, Median [IQR] | 143.07 [112.48-190.51] | 151.72 [144.65-176.9] | 136.5 [107.88-195.83] | 141.9 [118.78-188.27] | 0.412 |  |
| SpO2, Median [IQR] | 96.53 [95.14-97.85] | 95.75 [93.31-98.14] | 96.59 [94.52-97.6] | 96.82 [95.31-97.89] | 0.357 |  |
| Driving pressure, Median [IQR] | 12.37 [9.83-14.76] | 12.58 [10.87-15.75] | 13.0 [8.67-14.88] | 12.0 [9.64-14.25] | 0.613 |  |
| Minute ventilation, L, Median [IQR] | 9.17 [8.26-10.87] | 9.52 [8.73-10.72] | 9.01 [8.3-10.99] | 9.18 [7.99-10.73] | 0.966 |  |
| PCO2, mmHg, Median [IQR] | 42.21 [37.95-46.88] | 38.82 [33.89-44.24] | 41.48 [38.0-46.16] | 43.9 [38.44-48.05] | 0.220 |  |
| PEEP, Median [IQR] | 11.65 [9.62-13.38] | 10.64 [8.5-12.04] | 12.0 [9.33-13.25] | 11.86 [9.86-13.48] | 0.607 |  |
| PH, Mean [CI] | 7.34 [7.33,7.36] | 7.37 [7.33,7.41] | 7.38 [7.36,7.4] | 7.31 [7.29,7.33] | <0.001<sup>**§</sup> | Severe vs Mild, Intermediate |
| PIP, Median [IQR] | 27.9 [24.67-31.0] | 26.81 [25.0-28.5] | 28.0 [25.13-31.45] | 28.0 [24.73-31.0] | 0.377 |  |
| Plateau pressure, Median [IQR] | 24.0 [19.75-27.0] | 24.83 [23.38-25.75] | 24.0 [21.5-28.0] | 24.0 [18.33-25.75] | 0.555 |  |
| Static compliance, Median [IQR] | 28.0 [23.24-35.22] | 28.88 [26.49-36.14] | 26.43 [22.58-32.14] | 29.68 [23.88-36.52] | 0.312 |  |
| Tidal PBW ratio, Median [IQR] | 6.51 [6.04-7.08] | 6.6 [6.1-7.03] | 6.52 [6.29-7.48] | 6.41 [5.94-6.6] | 0.126 |  |
| Tidal volume, mL, Median [IQR] | 400.0 [370.0-440.62] | 400.0 [385.0-417.08] | 400.0 [370.0-450.0] | 400.0 [350.0-425.0] | 0.758 |  |
| Ventilator ratio, Median [IQR] | 1.83 [1.56-2.01] | 1.71 [1.3-1.94] | 1.87 [1.56-2.0] | 1.83 [1.62-2.02] | 0.451 |  |

<sup>†</sup> p-value calculated by analysis of variance (ANOVA)/Kruskal–Wallis test
<sup>\*\*</sup> False discovery rate corrected p-value < 0.05
<sup>§</sup> Age adjusted (analysis of covariance [ANCOVA]) p-value < 0.05
Abbreviations: ALT=Alanine aminotransferase, AST=Aspartate aminotransferase, CI=confidence interval, CK=Creatine kinase, CRP=C-reactive protein, ESR=Erythrocyte sedimentation rate, FiO2=fraction of inspired oxygen, GCS=Glasgow Coma Scale, IQR=Interquartile range, LDH=Lactate dehydrogenase, MAP=Mean arterial pressure, PaO2=partial pressure of oxygen, PBW=predicted body weight, PCO2=arterial partial pressure of carbon dioxide, PEEP=Positive end-expiratory pressure, PIP=Peak inspiratory
pressure, P/F ratio= $\text{PaO}_2/\text{FiO}_2$ ratio, $\text{SpO}_2$ =oxygen saturation.

**Table E-7.**
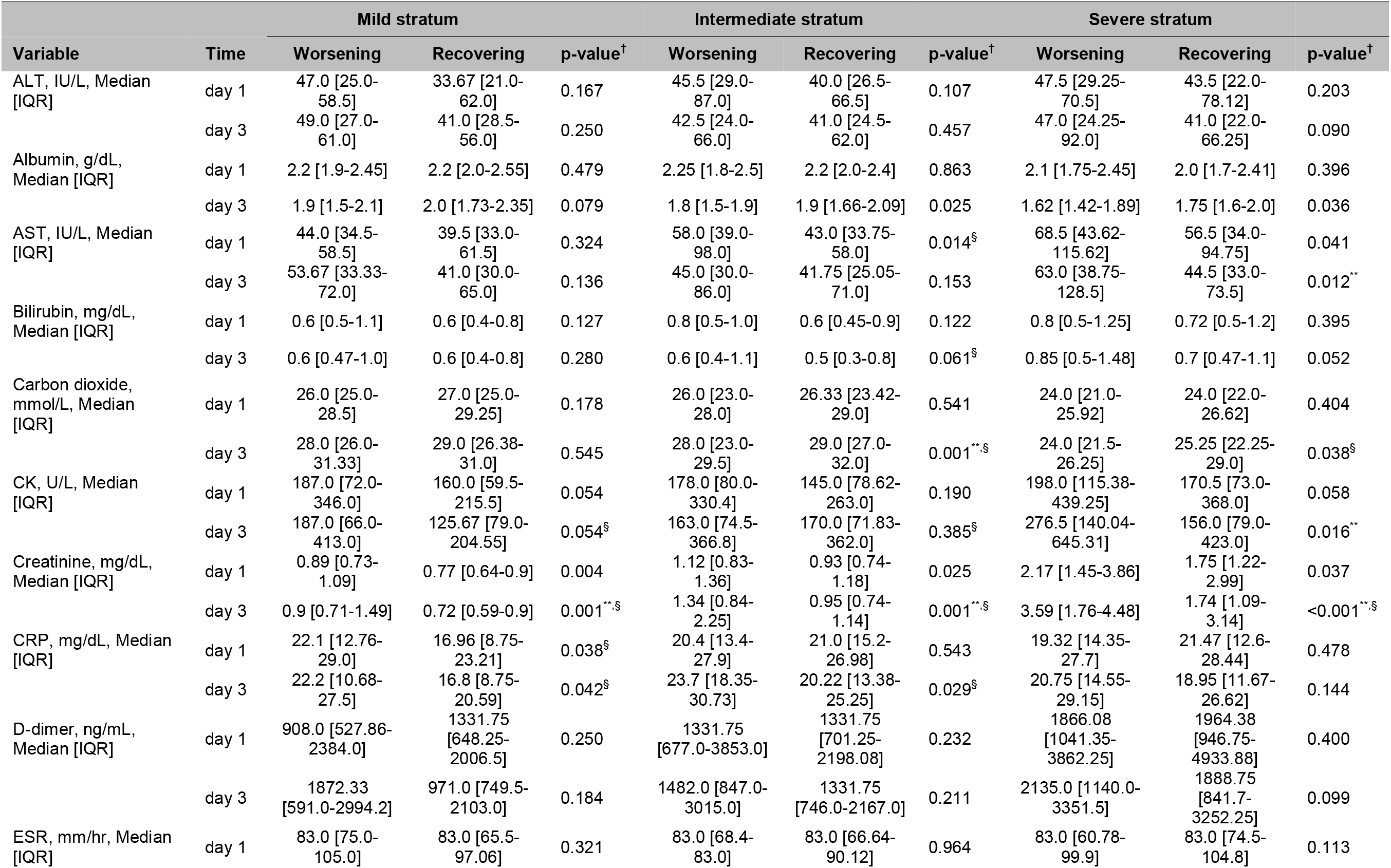
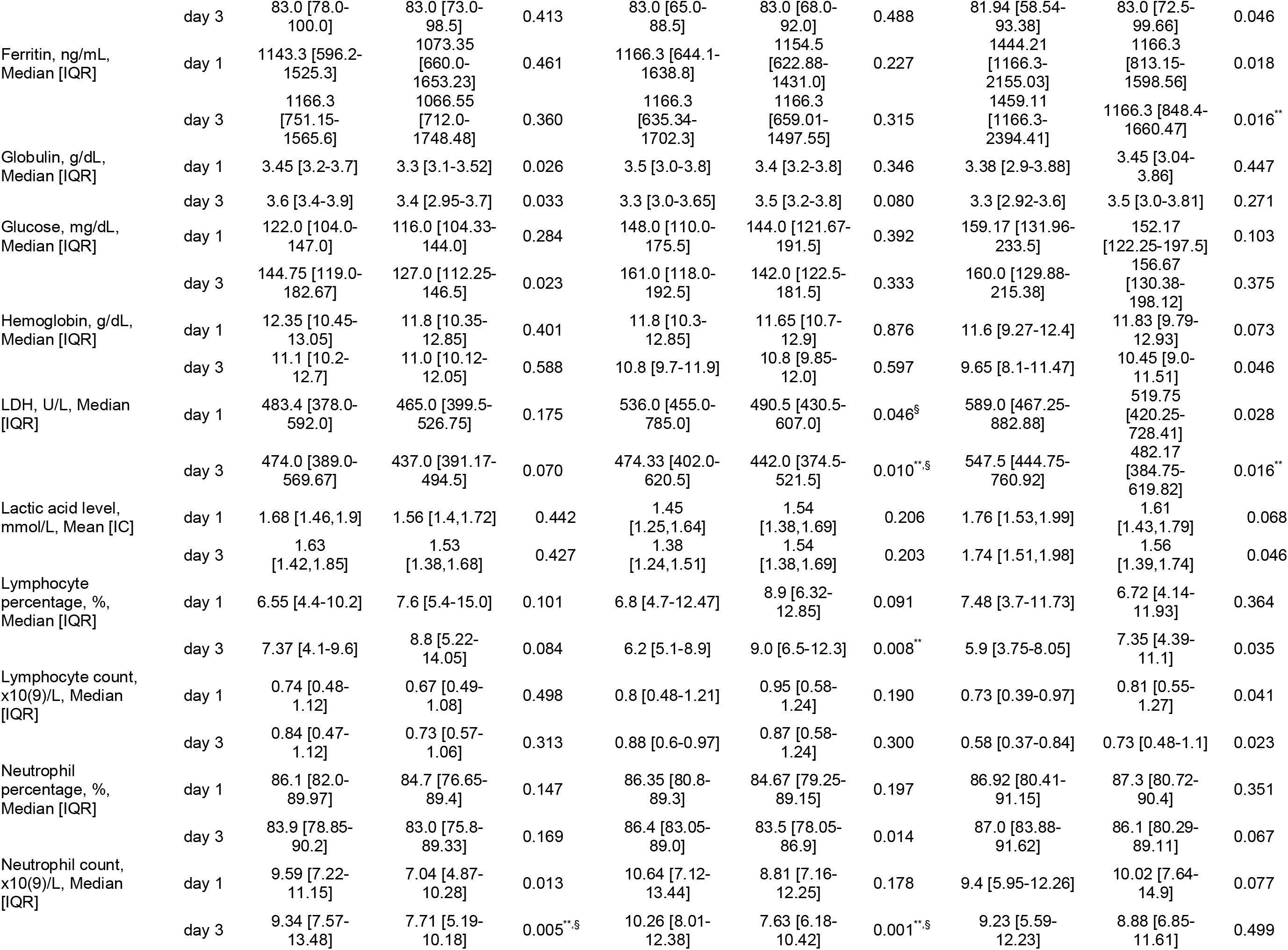
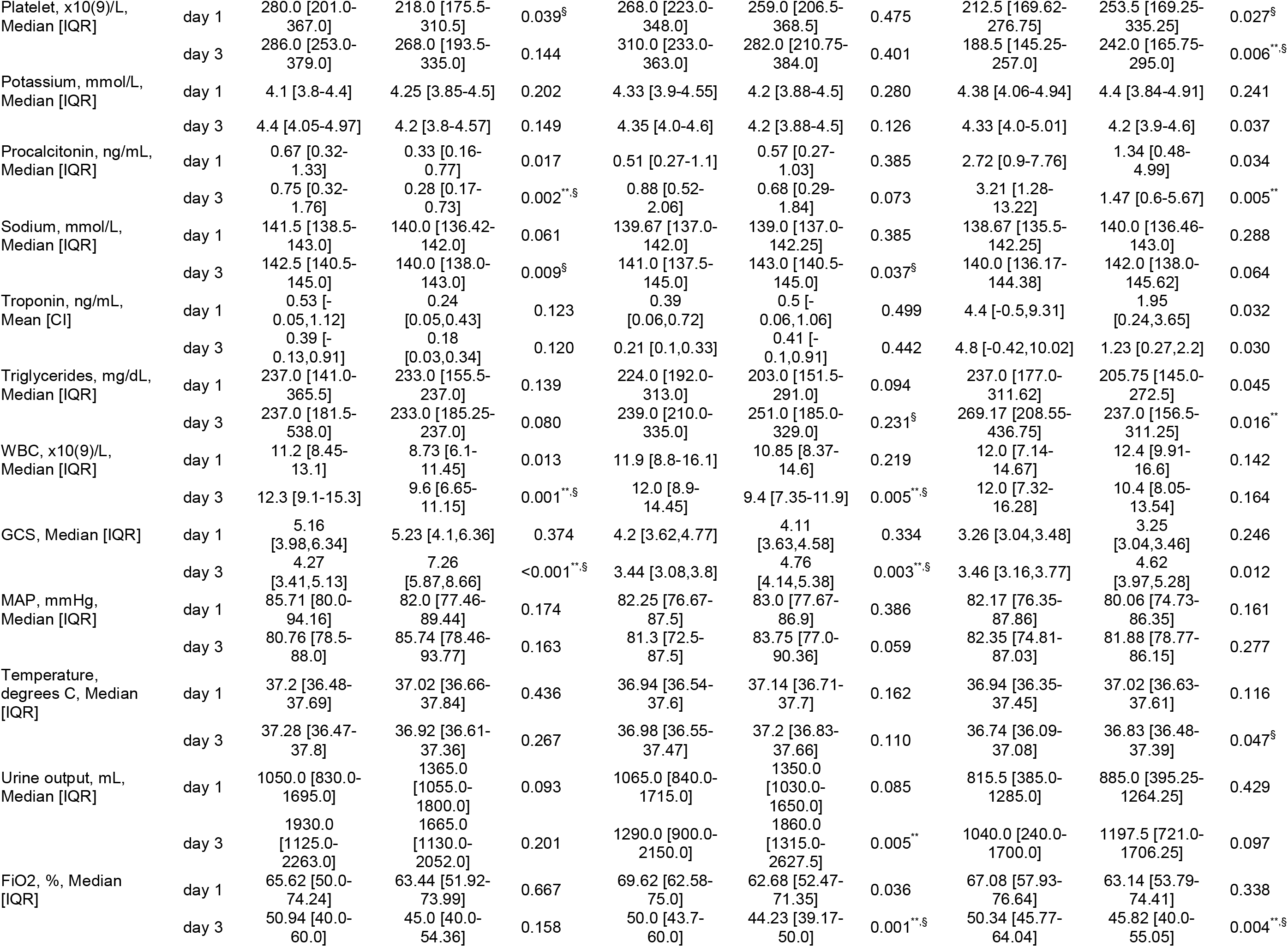
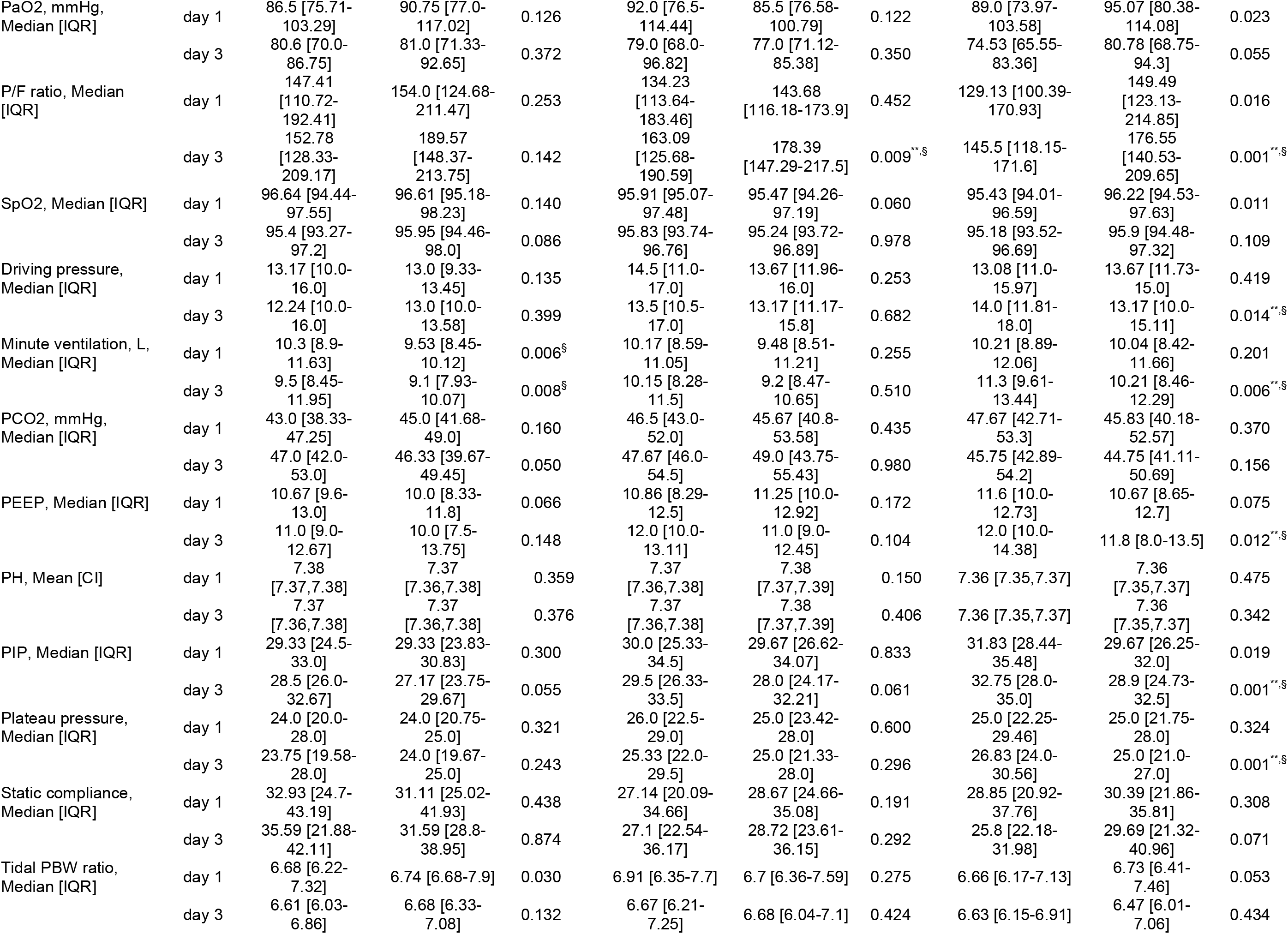
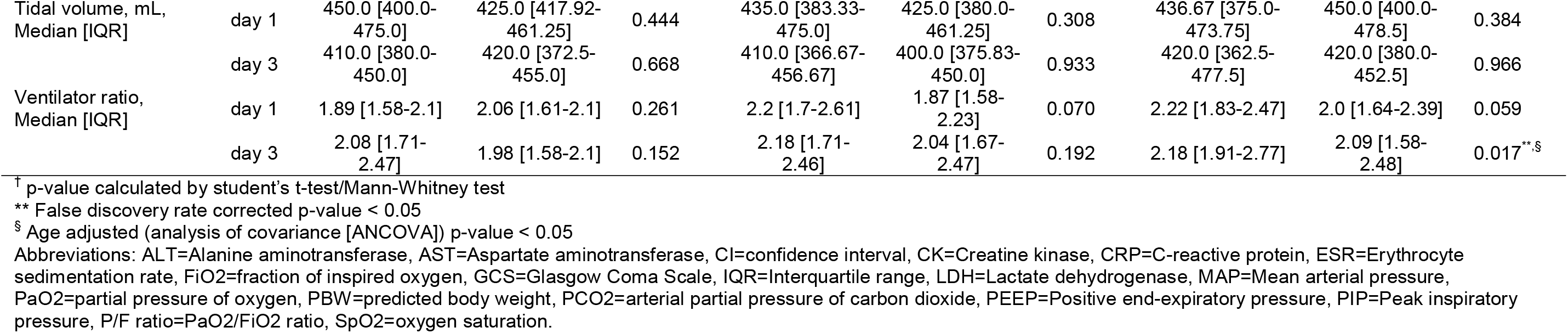
Clinical variables (laboratory test results, vital signs, respiratory variables, and ventilator parameters) of the trajectory subphenotypes in NYP-WCMC cohort. Data were examined at day 1 and day 3 post-intubation.

**Table E-8.**
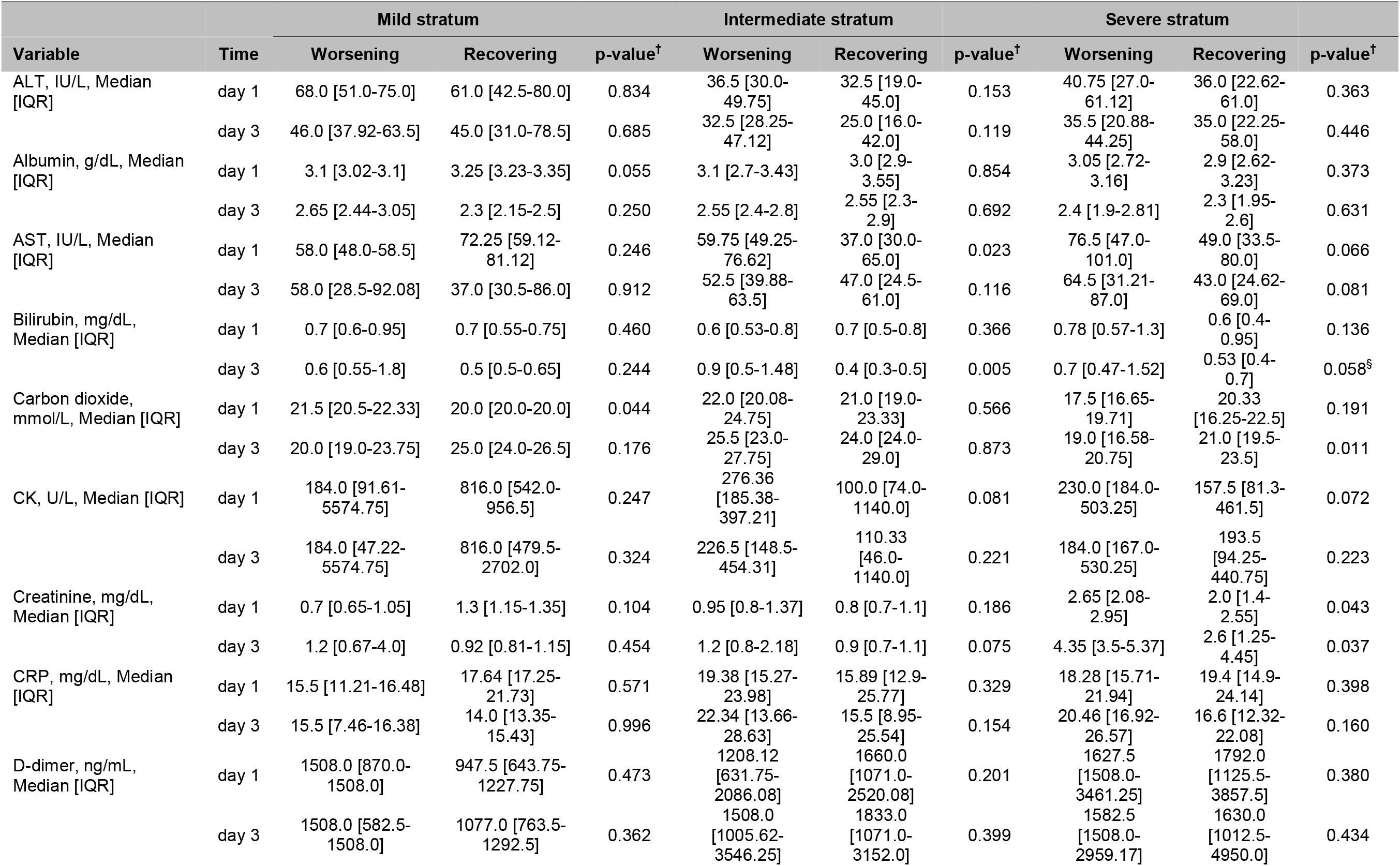
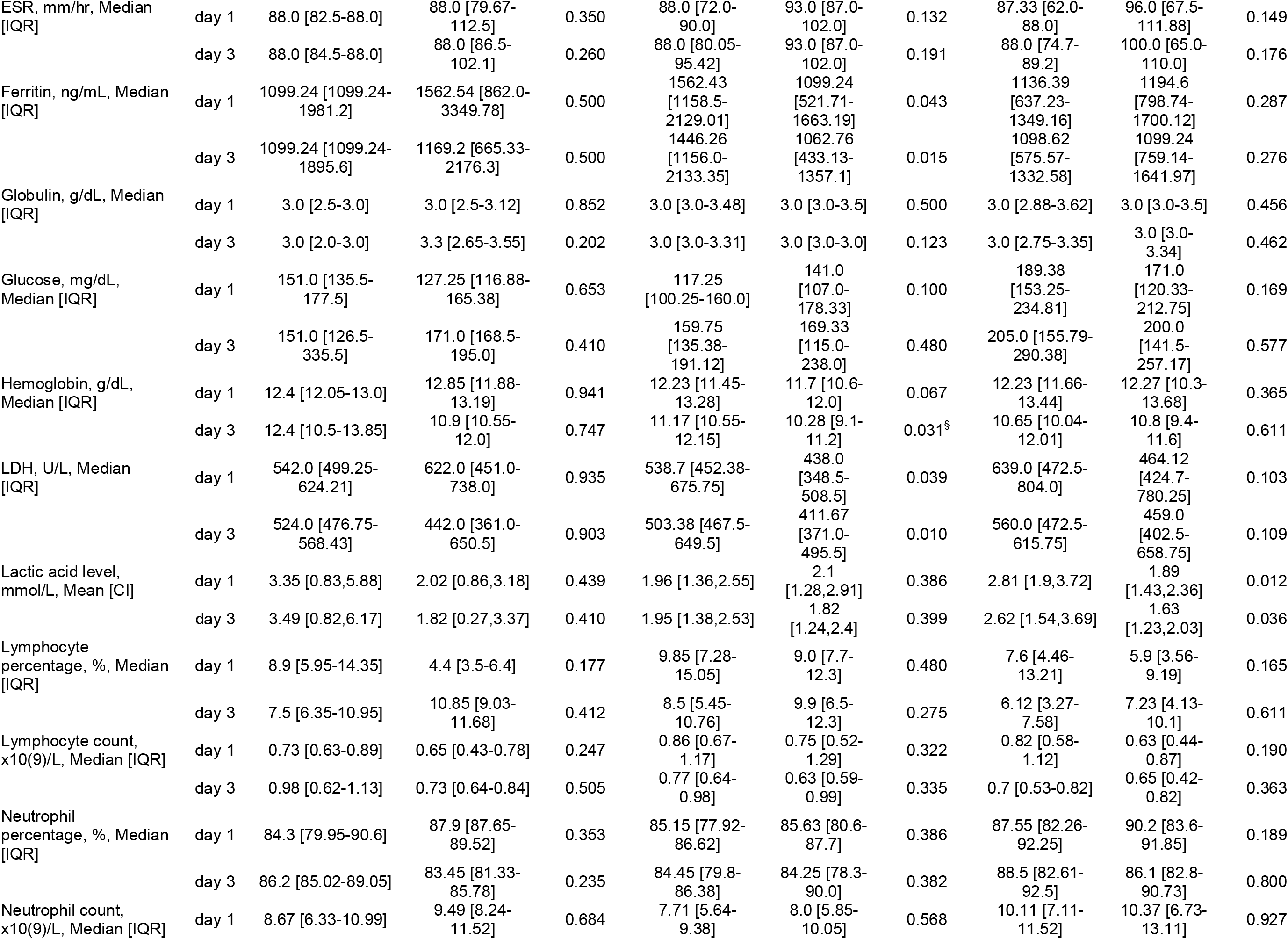
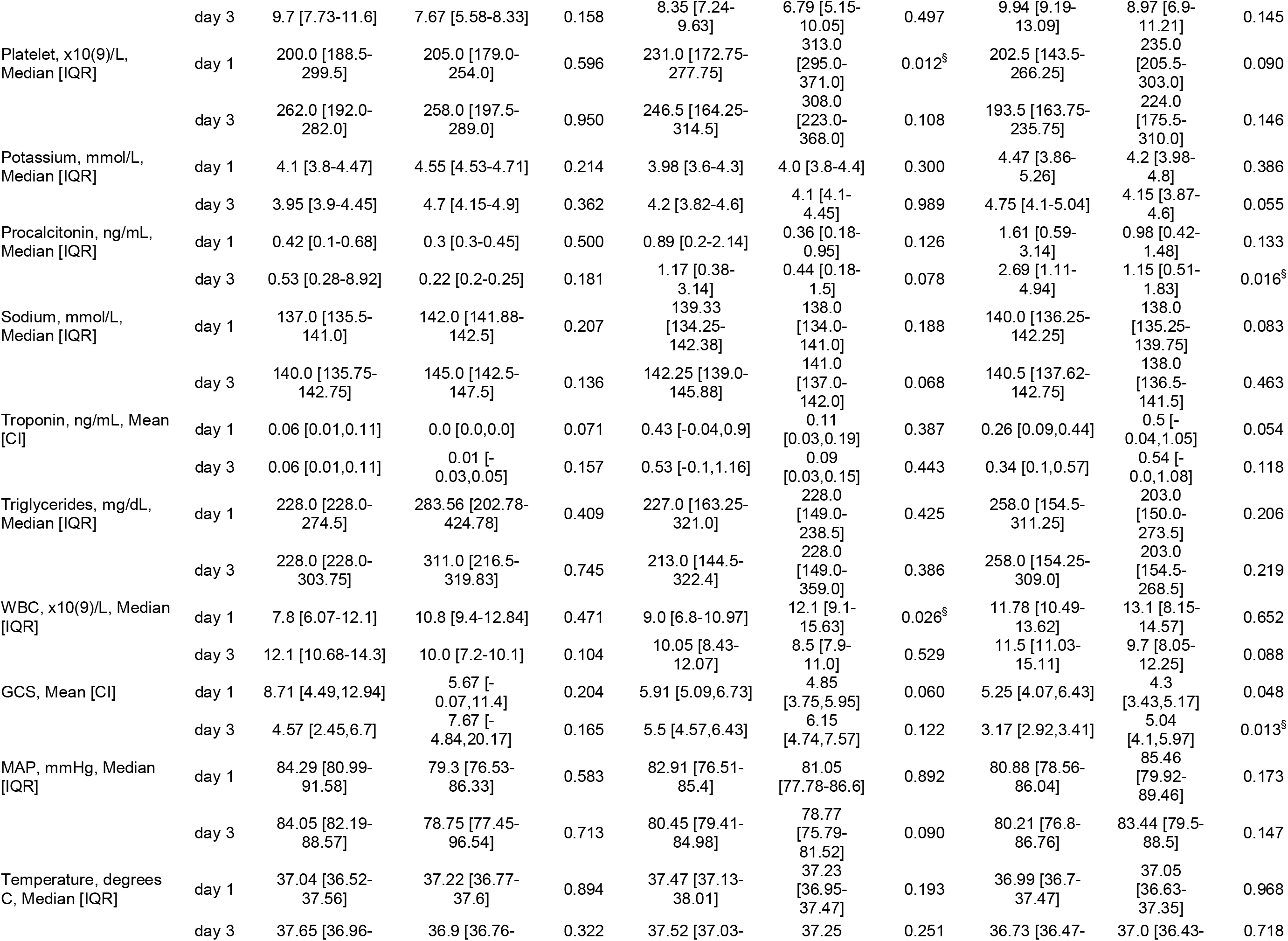
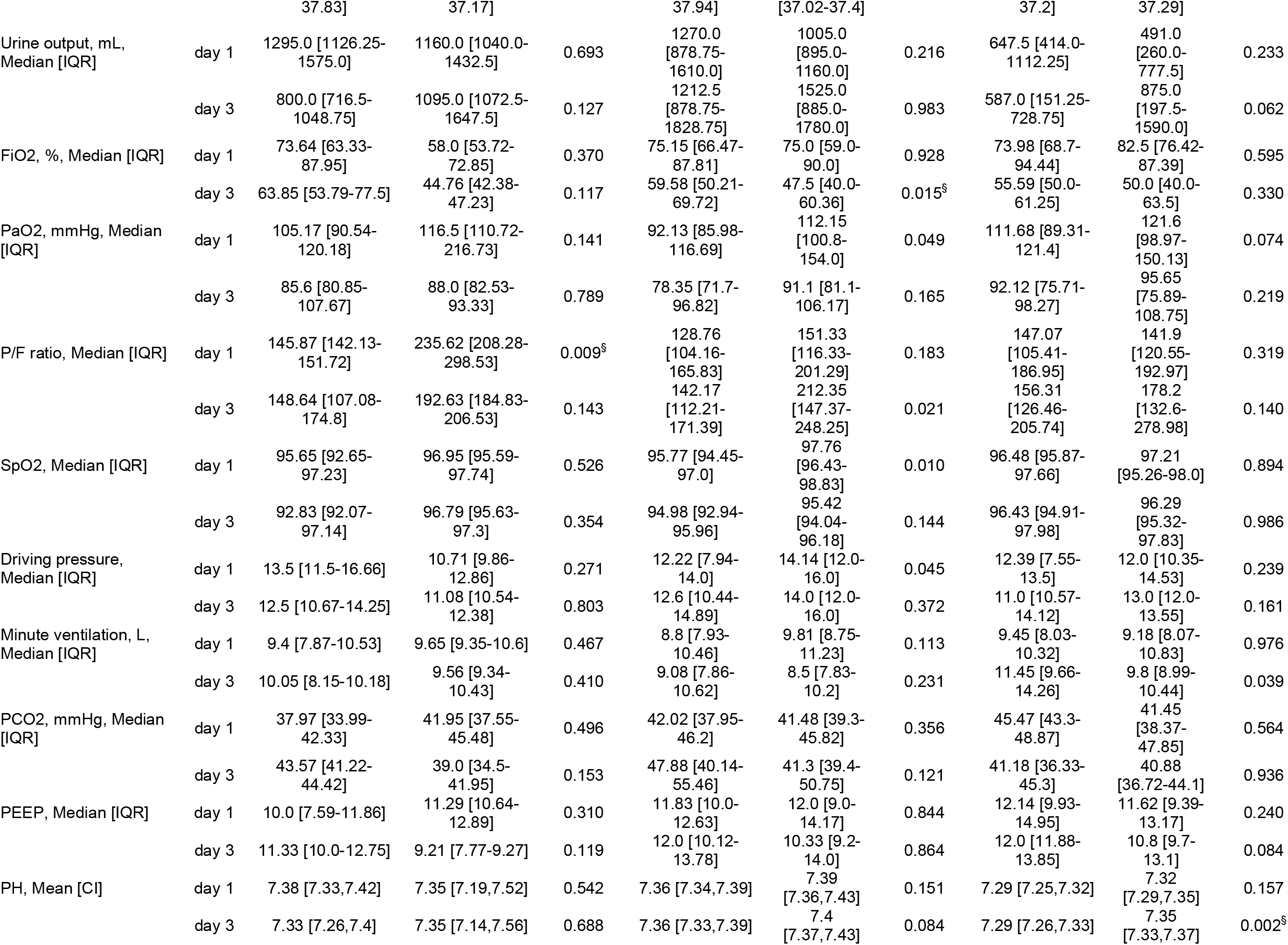
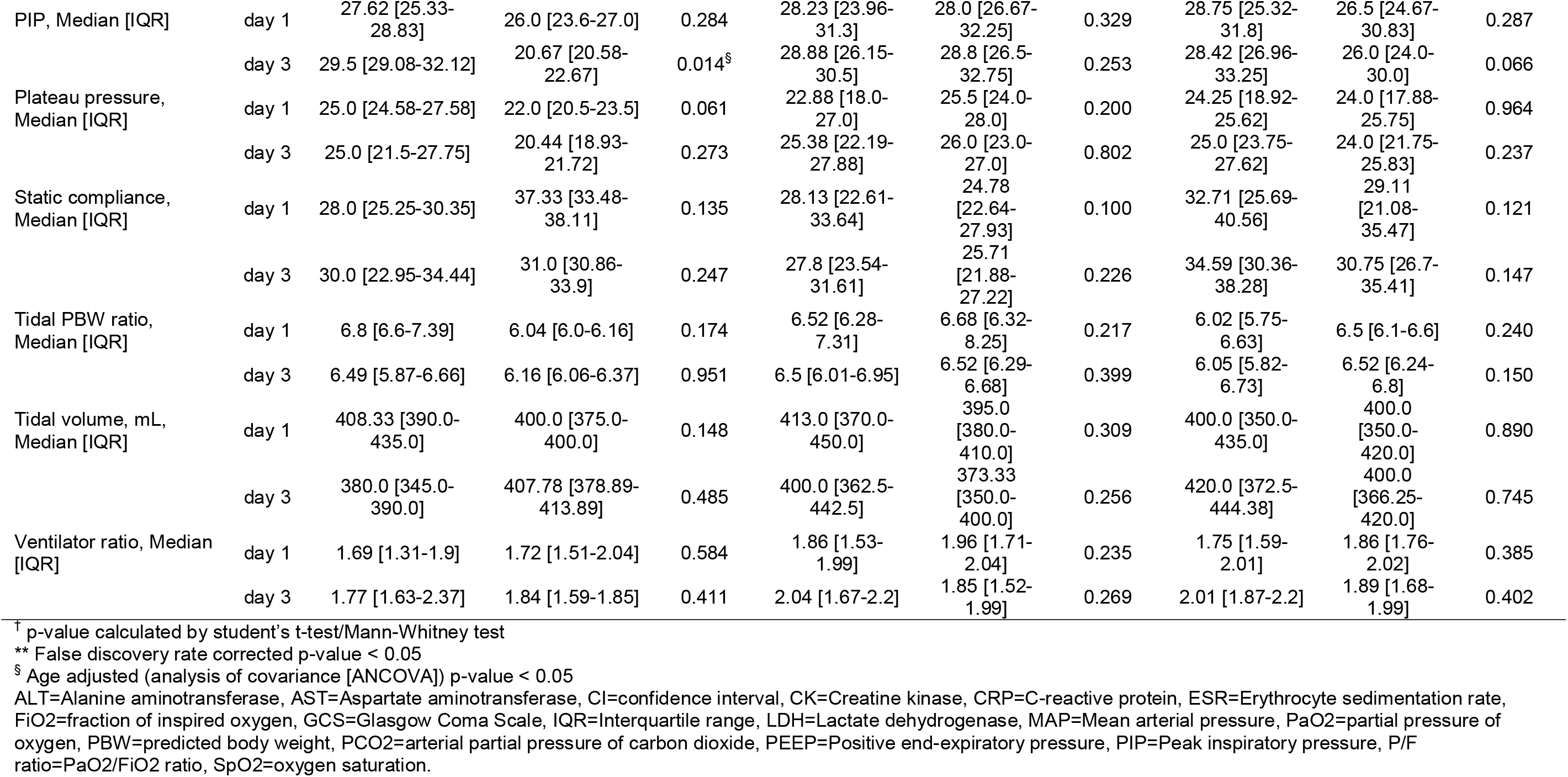
Clinical variables (laboratory test results, vital signs, respiratory variables) of the trajectory subphenotypes in NYP-LMH cohort. Data were examined at day 1 and day 3 post-intubation.

**Table E-9.**
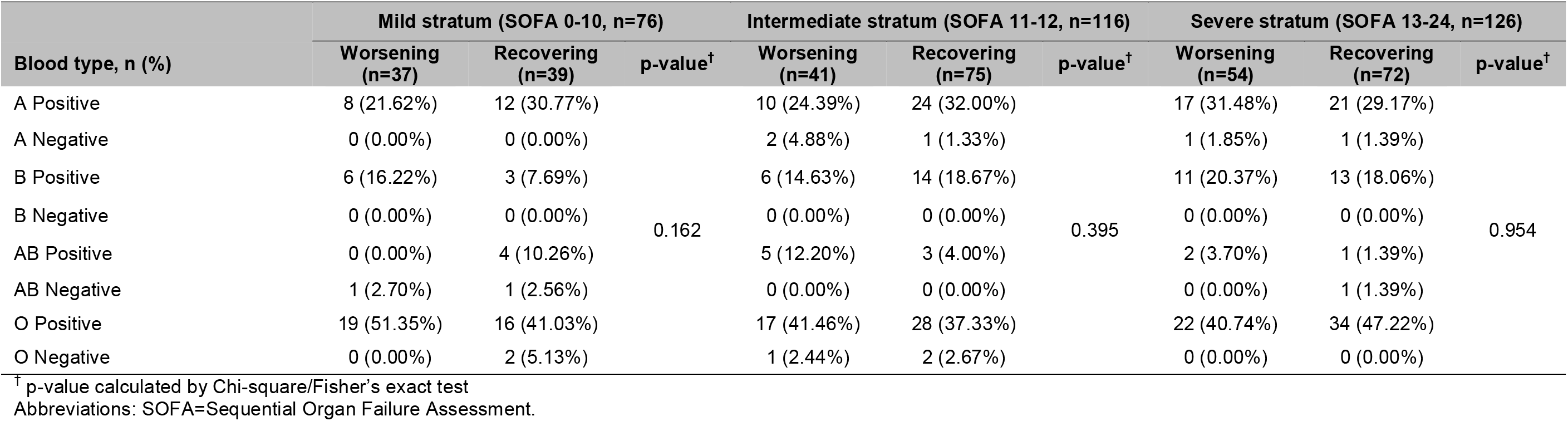
Blood type distribution of the trajectory subphenotypes in NYP-WCMC cohort.

**Table E-10.**
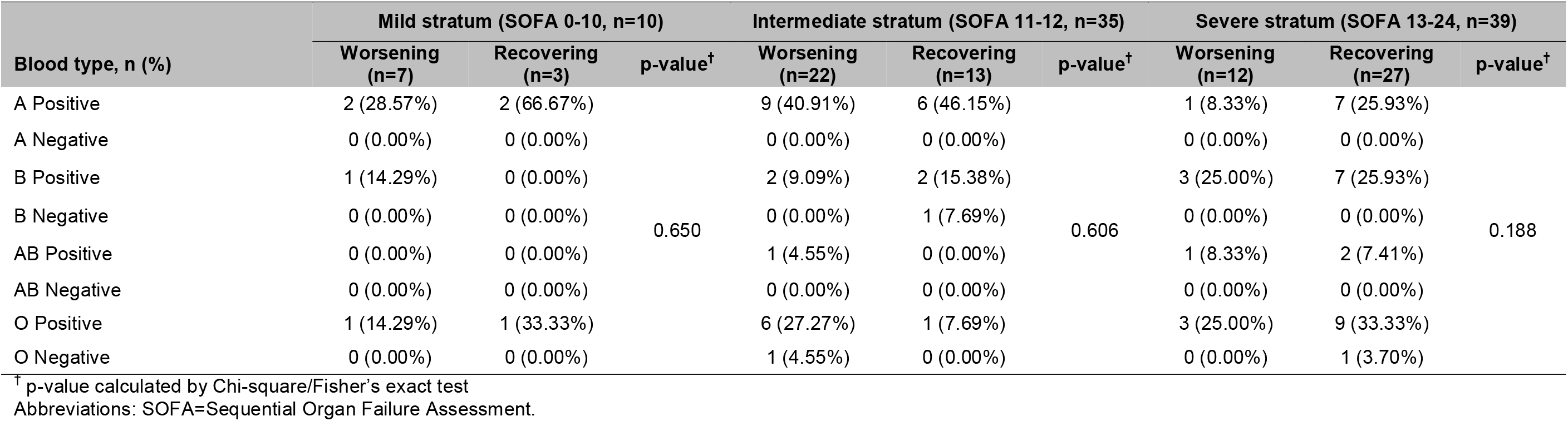
Blood type distribution of the trajectory subphenotypes in NYP-LMH cohort.

## Figures

**Figure E-1.**
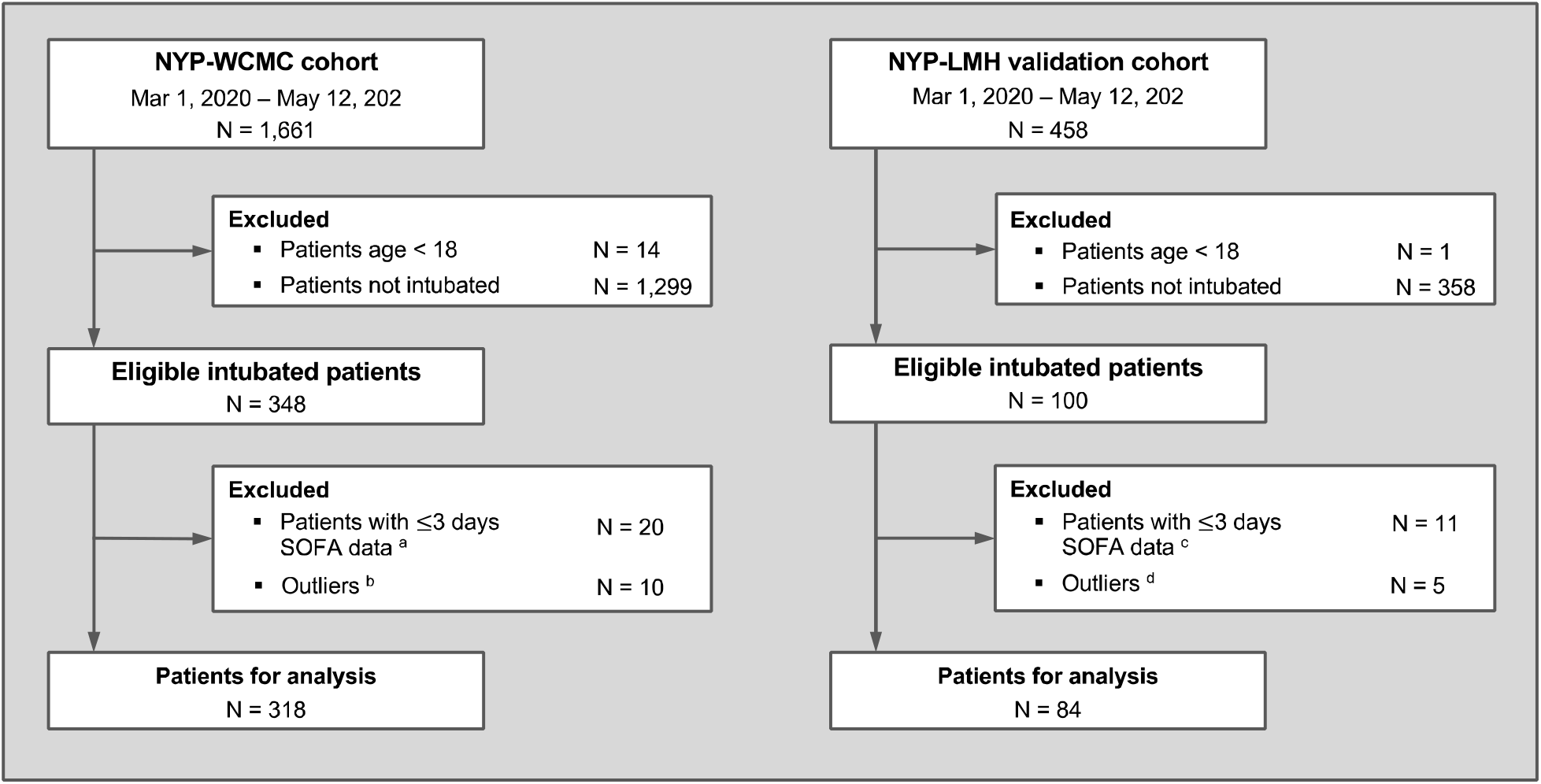
Patient exclusion criteria. Abbreviations: SOFA=Sequential Organ Failure Assessment. ^a^ Out of the 20 patients, 9, 3, and 3 dead within day 1, 2, 3 after intubation, respectively; 2 have no records, 2 and 1 only have 2- and 3-days SOFA data, respectively. ^b^ Out of the 10 patients, 7 have no change of SOFA score within 7 days after intubation, 3 whose SOFA trajectories fluctuated heavily. ^c^ Out of the 11 patients, 2, 5, and 4 dead within day 1, 2, 3 after intubation, respectively. ^d^ SOFA trajectories of the 5 patients fluctuated heavily. Abbreviation: NYP-LMH=New York Presbyterian Hospital-Lower Manhattan Hospital, SD=standard deviation, SOFA=Sequential Organ Failure Assessment

**Figure E-2.**
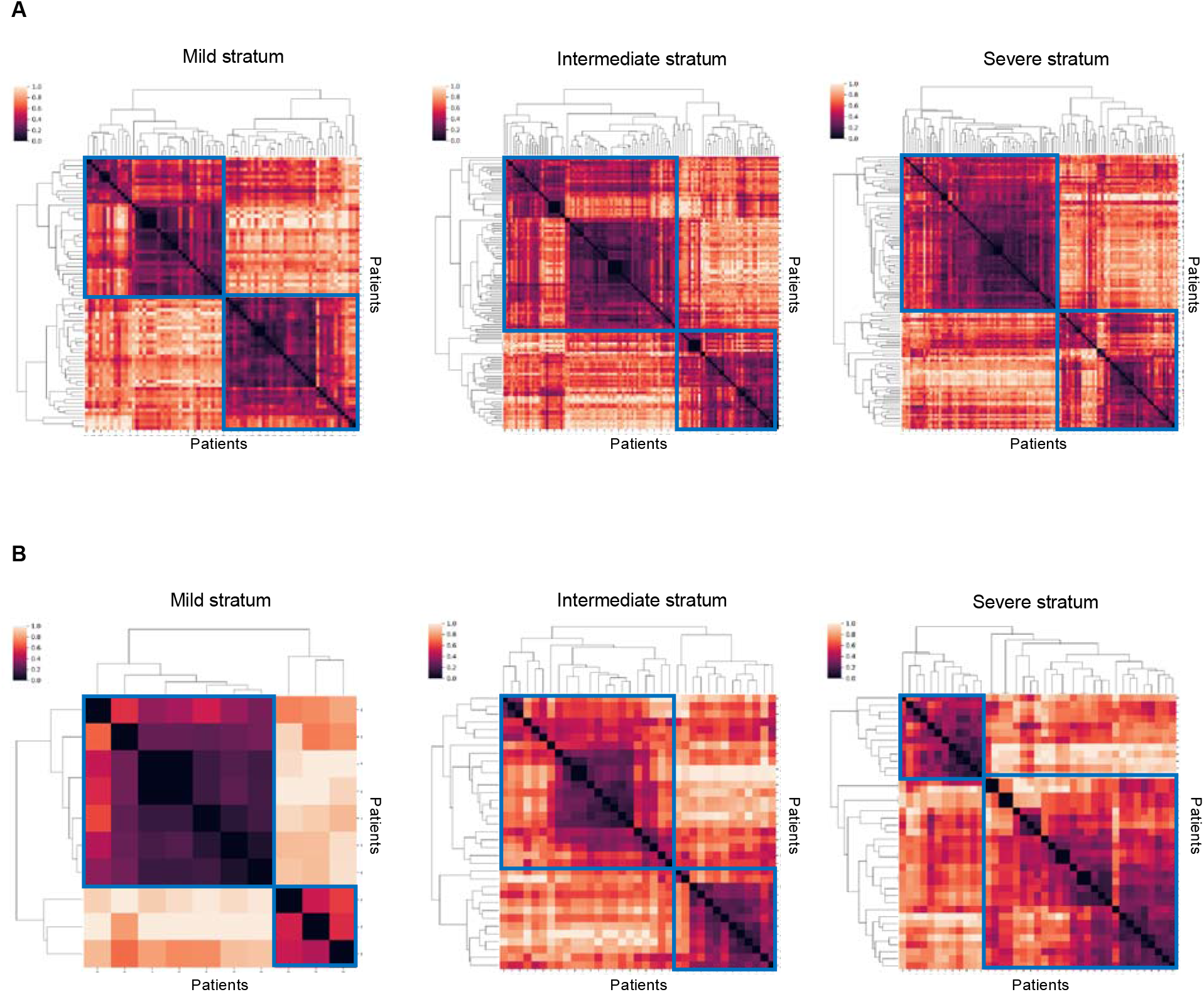
Clustergrams of hierarchical clustering. Horizontal and vertical axes represent patients. Color intensity denotes normalized pairwise patient similarity derived using Dynamic Time Warping (DTW). All clustergrams suggest optimal cluster number 2. *(A)* Clustergrams derived from the NYP-WCMC cohort. *(B)* Clustergrams derived from the NYP-LMH validation cohort.

**Figure E-3.**
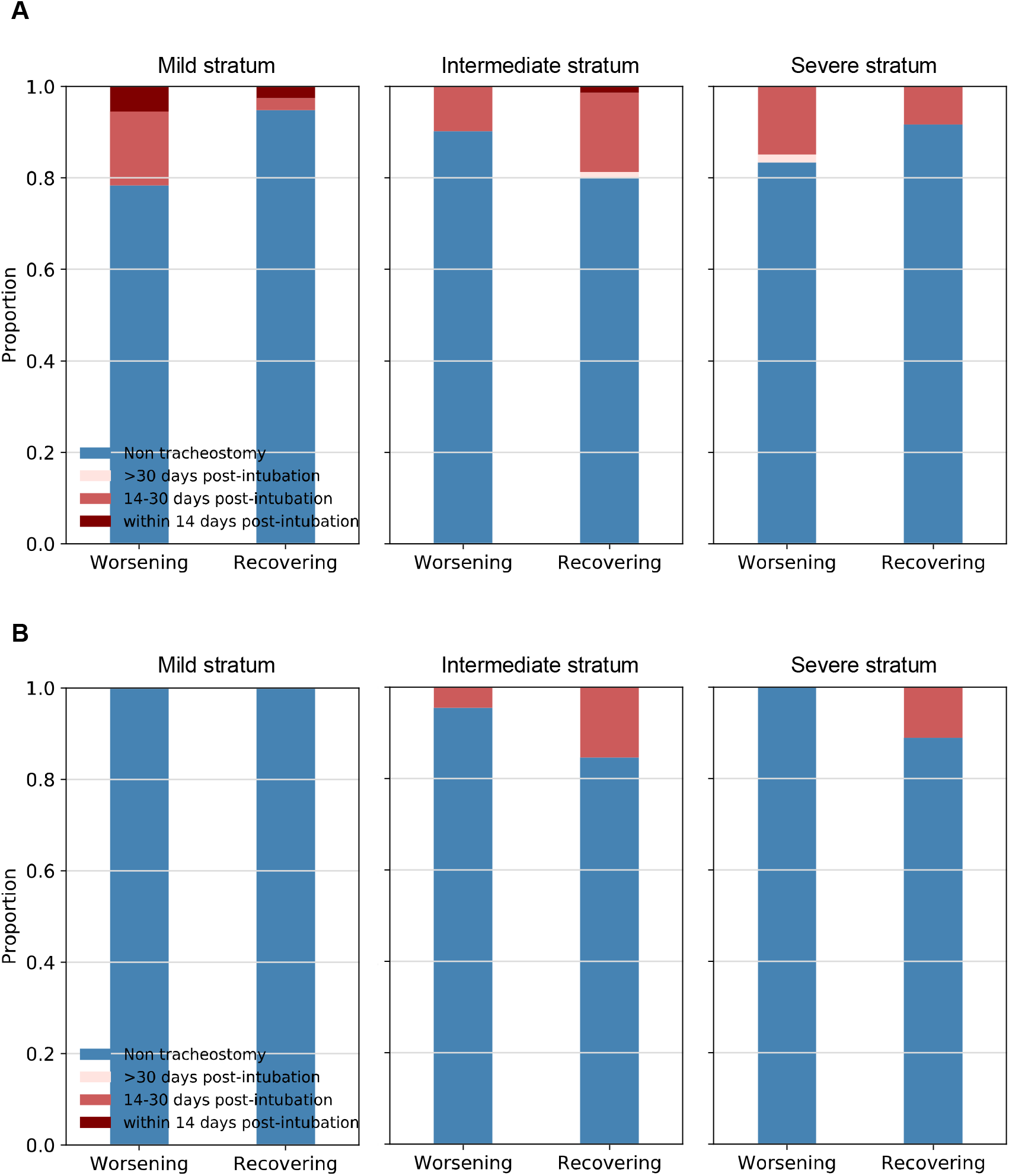
Tracheostomy outcome of the trajectory subphenotypes. (*A*) Statistics of tracheostomy of subphenotypes within the NYP-WCM cohort; (*B*) Statistics of tracheostomy of subphenotypes within the NYP-LMH validation cohort.

**Figure E-4.**
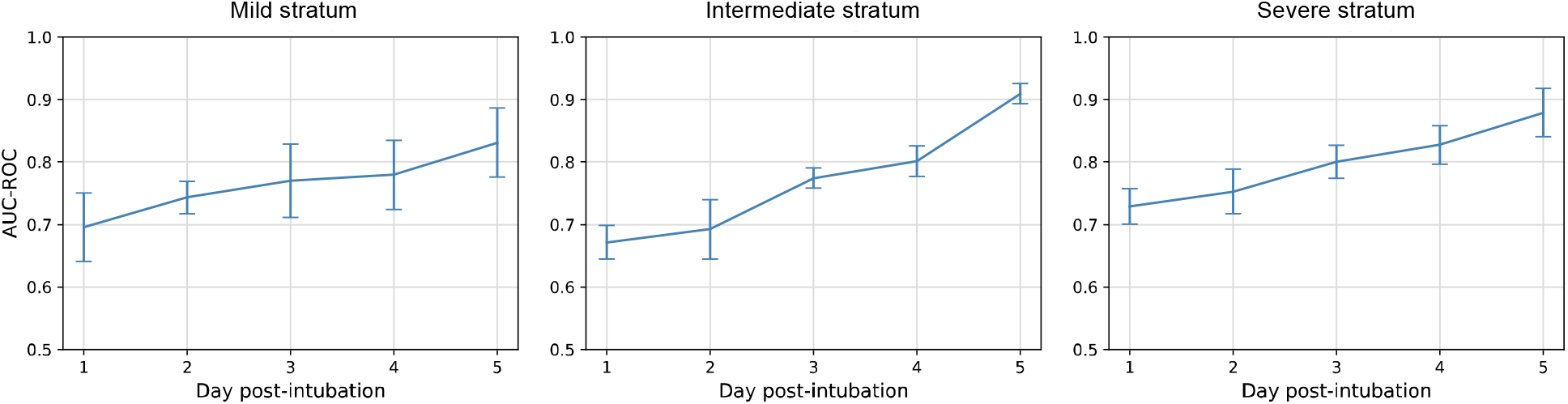
Area Under the Receiver Operating Characteristics (AUC-ROC) of subphenotype prediction models within baseline mild, intermediate, and severe strata. An AUC-ROC measures accuracy of a prediction model by comprehensively considering true positive rate and false positive rate in prediction. Abbreviations: AUC-ROC=Area Under the Receiver Operating Characteristics.

**Figure E-5.**
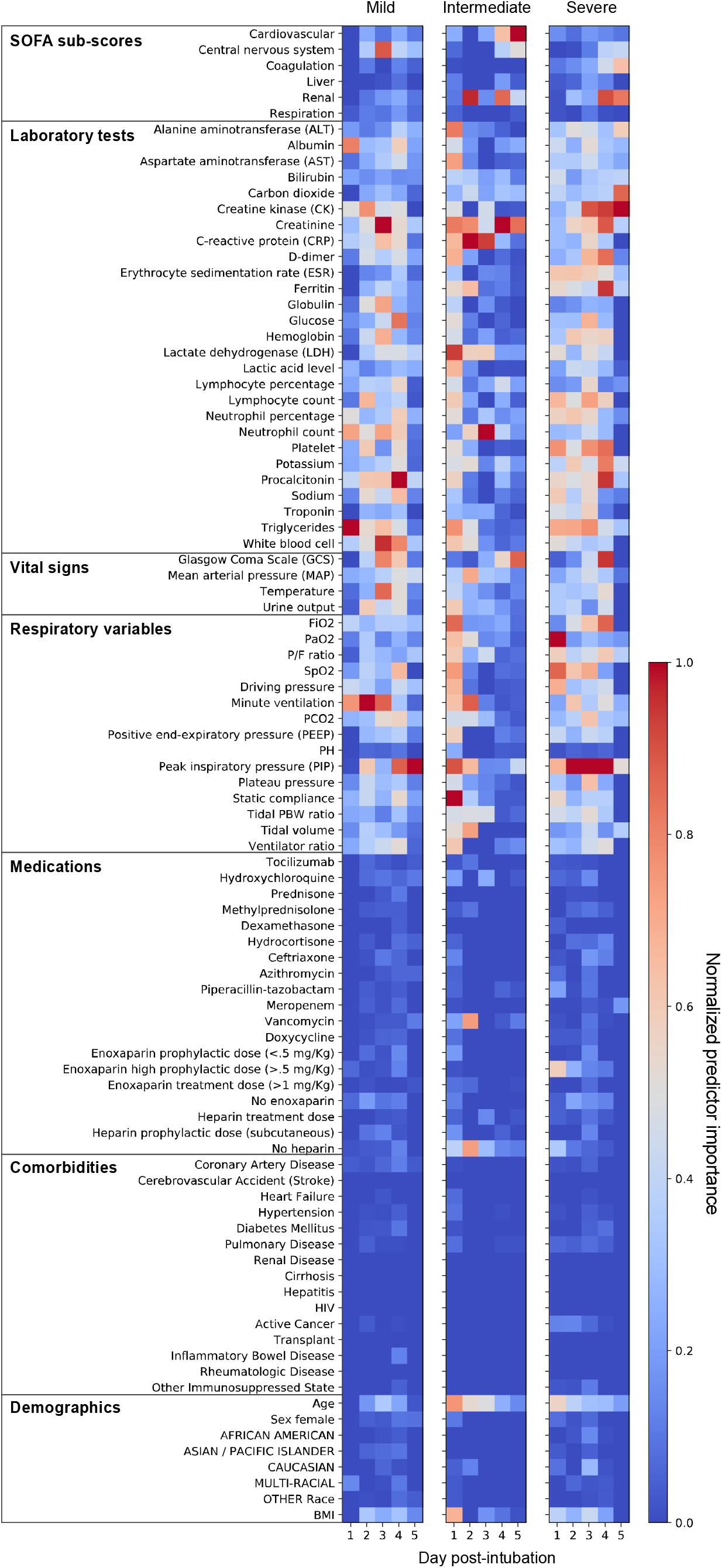
Predictor importance of the subphenotype prediction models. Horizontal axis of each heatmap presents timepoint post-intubation when data were used to train the random forest-based prediction model of the worsening and recovering subphenotypes. Color intensity represents the normalized importance of a predictor in a specific prediction model. Abbreviations: BMI=body mass index, FiO2=fraction of inspired oxygen, HIV=human immunodeficiency viruses, PaO2=partial pressure of oxygen, PBW=predicted body weight, PCO2=arterial partial pressure of carbon dioxide, P/F ratio=PaO2/FiO2 ratio, SpO2=oxygen saturation.

## Notes

**Funding:** This study received support from NewYork-Presbyterian Hospital (NYPH) and Weill Cornell Medical College (WCMC), including the Clinical and Translational Science Center (CTSC) (UL1 TR000457) and Joint Clinical Trials Office (JCTO). FW and CS are supported by NSF IIS 2027970, 1750326, ONR N00014-18-1-2585.

### Competing Interest Statement

The authors have declared no competing interest.

### Clinical Trial

n/a

### Funding Statement

This study received support from New York-Presbyterian Hospital (NYPH) and Weill Cornell Medical College (WCMC), including the Clinical and Translational Science Center (CTSC) (UL1 TR000457) and Joint Clinical Trials Office (JCTO). FW and CS are supported by NSF IIS 2027970, 1750326, ONR N00014-18-1-2585.

### Author Declarations

The study is approved by the IRB of Weill Cornell Medicine with protocol number 20-04021909.

